# Wearable-based sleep and activity tracking detects Parkinson’s disease and associated pathological hallmarks

**DOI:** 10.64898/2026.11.13.26367931

**Authors:** Apostolos Moustaklis, Ioannis Gerasimou, Charalampos Sotirakis, Paraskevi Chairta, Monica M. Kurtis, Natalia Del Campo, Tim Feige, Nils Schnalke, Maria-Luisa Almarcha Menargues, Kyriaki Michailidou, Kyproula Christodoulou, Margherita Fabbri, Björn H. Falkenburger, Leontios Hadjileontiadis, Stelios Hadjidimitriou

## Abstract

Parkinson’s disease (PD) is associated with sleep and motor function impairment, even in the early stages. Here, we investigated whether passive wearable-based sleep and activity tracking can detect PD and its associated pathological hallmarks, dopaminergic deficit and α-synuclein pathology. Sleep and activity metrics (SAM) were derived from free-living wrist accelerometry data of a U.S. cohort and used to train a machine learning model (SAM-PD) to distinguish persons with PD (n = 133) from healthy controls (n = 26). The model achieved an AUROC of 0.93 and an AUPRC of 0.98, demonstrated excellent test-retest reliability, and generalized without retraining to an independent European cohort (n = 40; 50% PwPD) monitored with a different device (AUROC, 0.95; AUPRC, 0.95). REM sleep duration, walking cadence, and low-level acceleration features were most predictive. Applied to a U.S. at-risk cohort, SAM-PD performed comparably to the resource-intensive Movement Disorder Society (MDS) prodromal PD risk score in detecting dopaminergic deficit (AUROC, 0.86 vs. 0.80; AUPRC, 0.45 vs. 0.46; n = 121; 12.4% prevalence) and α-synuclein pathology (AUROC, 0.76 for both; AUPRC 0.50 vs. 0.42; n = 114; 16.7% prevalence). A combination with the MDS score modestly improved detection. Findings support further evaluation of wearable monitoring for PD screening and biomarker enrichment in larger populations.

## Introduction

Parkinson’s disease (PD) is a progressive neurodegenerative disorder defined by two primary, connected pathological hallmarks: the loss of dopaminergic neurons in the brain’s nigrostriatal pathway and the accumulation of misfolded α-synuclein proteins in the central and peripheral nervous systems [1]. The disease manifests with cardinal motor symptoms such as resting tremor, bradykinesia, and rigidity, as well as early non-motor signs including, *inter alia*, sleep disturbances, olfactory loss, and depression. It is estimated that by the time of clinical diagnosis, more than 50% of dopaminergic neurons have been lost [2]. The prodromal phase of PD may therefore begin years before the onset of the key motor features [3]. This window presents an opportunity for effective intervention and testing of neuroprotective and disease-modifying therapies, rendering early detection crucial.

Dopamine transporter (DAT) single-photon emission computed tomography (SPECT) scanning, also referred to as DAT scan, has shown high diagnostic precision for identifying nigrostriatal dopaminergic neurodegeneration and is frequently employed as a surrogate endpoint for PD progression in prodromal research [4]. More recently, cerebrospinal fluid (CSF)-based α-synuclein seed amplification assays (SAA) have been shown to provide a more direct etiologic confirmation of the disease’s underlying synucleinopathy through the detection of misfolded α-synuclein aggregates [5, 6]. Although reliable, both tests require specialized clinical resources that limit their suitability for cost-efficient large-scale screening. Research criteria based on risk factors and the emergence of non-motor and early motor symptoms have also been proposed for diagnosing prodromal PD [7, 8]. While useful, their application requires neurological evaluations and niche ancillary testing, potentially limiting the identification of individuals with early, subclinical disease [9]. Therefore, an accessible, unobtrusive, and scalable method remains essential for identifying individuals at risk of PD who warrant specialist assessment and confirmatory biomarker testing.

With their growing adoption, mobile and wearable technologies offer promising opportunities. Recent work has demonstrated the potential of fit-for-purpose smartphone-based tasks to predict DAT scan results [10], yet, such approaches require active engagement, which may limit adherence [11], and may be susceptible to behavioral changes due to monitoring awareness (the Hawthorne effect). In contrast, sleep and activity profiles, which are affected by PD even in its early stages owing to established prodromal conditions such as REM sleep behavior disorder (RBD) and sub-threshold parkinsonism [8, 12], can be passively tracked via wearable data [13, 14]. Latest studies demonstrate that relevant features derived from wrist wearables could identify PD years before clinical diagnosis [14] and are associated with dopaminergic deficit and α-synuclein pathology in individuals at risk of the disease [15]. Nevertheless, recent work showed that cross-cohort generalization of wearable metrics-based machine learning models remains a challenge [16]. For clinical translation, however, wearable-derived measures must produce consistent results across device hardware and populations, demonstrate temporal reliability, and rely on reproducible and interpretable algorithms [17–19].

This study investigates whether sleep and activity metrics (SAM) derived from passively collected free-living wrist-wearable data can identify persons with PD (PwPD), dopaminergic deficit, or α-synuclein pathology. Leveraging wrist accelerometry data from international cohorts, we developed the machine learning-based SAM-PD score and evaluated its performance in differentiating: (1) individuals with PD from healthy controls (HCs); (2) at-risk individuals with normal or abnormal DAT scan results; and (3) at-risk individuals with positive or negative CSF α-synuclein SAA results. We further compared the SAM-PD score against the Movement Disorder Society (MDS) prodromal PD risk score and investigated whether their combination enhances detection beyond what either approach achieves alone across several pathological hallmark scenarios. To ensure trustworthiness of the SAM-PD score, we assessed explainability, test-retest reliability, performance bias across key sensitive variables, and generalizability across different device hardware and geographic regions. With SAM-PD, we aim to introduce a digital tool that could complement established clinical measures and pave the way for unobtrusive, scalable, and cost-effective screening of individuals at risk of PD.

## Results

### Cohort characteristics

We analyzed free-living wrist accelerometry data from 323 participants in the U.S.-based Verily Study Watch cohort of the Parkinson’s Progression Markers Initiative (PPMI) study (NCT01141023) and 81 participants in the European AI-PROGNOSIS Digital Biomarkers Development (dBM-DEV) study (NCT06444789). Participants in the Verily Study Watch cohort were monitored for 484 ± 202 days (mean ± standard deviation) using the Verily Study Watch device (Verily Life Sciences LLC, Dallas, TX, USA), whereas participants in the dBM-DEV study were monitored for 40 ± 29 days using the Garmin Vivoactive 5 device (Garmin Ltd., Schaffhausen, Switzerland). After applying inclusion criteria requiring at least one 14-day monitoring window including seven days and seven nights of adequate wearable data (‘Datasets and cohorts’, Methods), 283 participants from the Verily Study Watch cohort (133 PwPD, 26 HCs, and 124 individuals at risk of PD) and 61 participants from the dBM-DEV study (20 PwPD, 20 HCs, and 21 individuals at risk of PD) qualified for further analysis. The demographic and clinical characteristics of included participants are summarized in Table 1. No statistical differences in sex, age, or body mass index (BMI) were observed between PwPD and HCs in either cohort (p ≥ 0.197). Within the Verily Study Watch cohort, at-risk participants differed in age from both HCs (p < 0.05) and PwPD (p < 0.001) and included a higher proportion of females than PwPD (61.3% vs. 40.6%, p < 0.001). A similar age difference was observed in the dBM-DEV cohort between at-risk participants and both HCs (p < 0.05) and PwPD (p < 0.001). Between cohorts, significant differences were observed in age among both PwPD (p < 0.001) and at-risk participants (p < 0.001), and in sex for the at-risk group only (p = 0.001). Detailed results of statistical comparisons are provided in Supplementary Table 1.

**Table 1.**
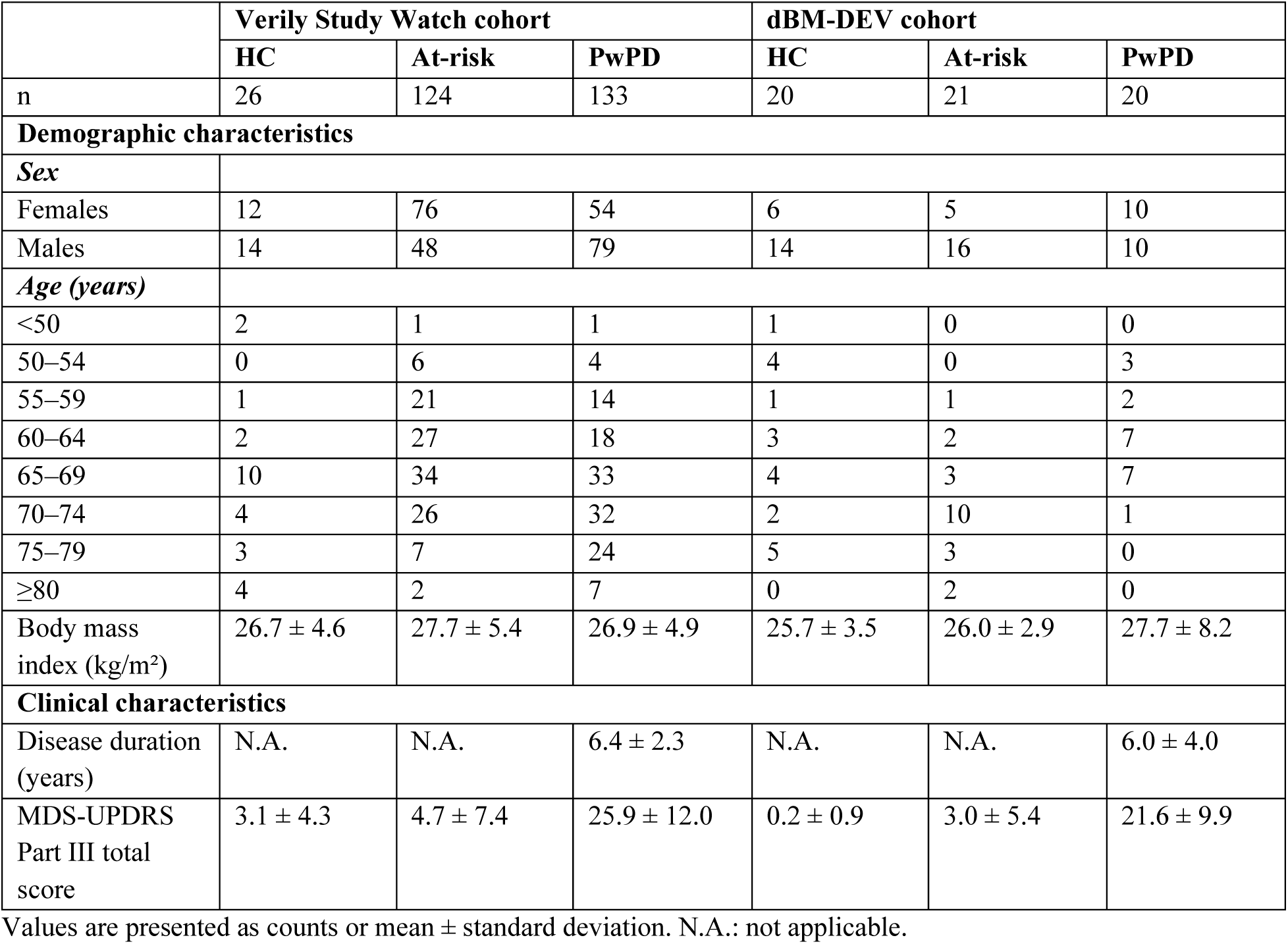
Demographic and clinical characteristics of study participants who qualified for analysis after applying the inclusion criteria for adequate wearable data.

For the Verily Study Watch cohort, DAT scan and CSF α-synuclein SAA data were available. DAT scan imaging and CSF collection were performed before wearable data collection began, whereas SAA testing was conducted on the stored CSF samples after completion of wearable data collection. For analyses involving DAT scan results, we excluded PwPD and at-risk individuals whose DAT scans were acquired more than three years before the start of wearable data collection, while retaining all HCs regardless of the DAT scan acquisition date. Table 2 summarizes test availability and the time intervals between the tests and wearable data collection. The presence of dopaminergic deficit and α-synuclein pathology was determined by an abnormal DAT scan result and a positive CSF α-synuclein SAA, respectively (‘Evaluation against pathological hallmarks’, Methods). Based on the availability and results of the two tests, we evaluated four scenarios defined by the presence of pathological hallmarks, with the corresponding group stratification illustrated in Fig. 1: (a) dopaminergic deficit, (b) α-synuclein pathology, (c) either pathological hallmark present, and (d) both pathological hallmarks present. In the PwPD group, 85 of 86 participants had an abnormal DAT scan (98.8%) and 105 of 122 had a positive SAA test (86.1%). Overall, 59 of 60 PwPD (98.3%) were positive for both dopaminergic deficit and α-synuclein pathology, whereas 131 of 132 (99.2%) were positive for at least one pathological feature. None of the HCs had an abnormal DAT scan or a positive SAA test. In the at-risk group, the prevalence of dopaminergic deficit and α-synuclein pathology was 12.4% (15 of 121 participants) and 16.7% (19 of 114 participants), respectively. Seven of 96 at-risk participants (7.3%) were positive for both dopaminergic deficit and α-synuclein pathology, whereas 27 of 116 (23.3%) were positive for at least one pathological feature.

**Fig. 1.**
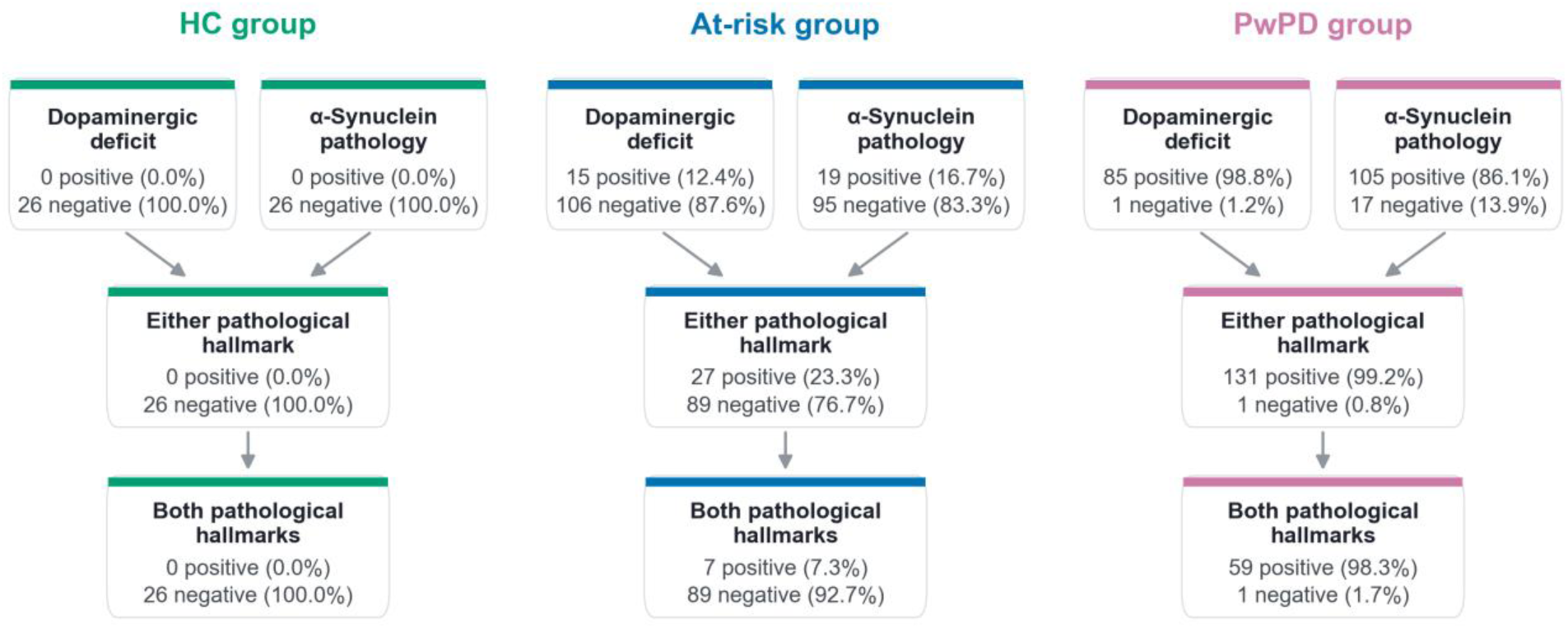
Group stratification according to pathological hallmark presence in the Verily Study Watch cohort. For each group (HC, At-risk, PwPD), the diagram shows the number of participants with eligible tests and the resulting counts and percentages of participants that were positive or negative for dopaminergic deficit and α-synuclein pathology, positive for either or both pathological hallmarks, or negative for both. Percentages are calculated relative to the total number of participants in each pathological hallmark scenario.

**Table 2.** Availability and timing of DAT scan and CSF α-synuclein SAA relative to wearable data collection in the Verily Study Watch cohort.

| **Test** |  | **HC** | **At-risk** | **PwPD** |
|---|---|---|---|---|
| DAT scan | n participants (Interval between DAT imaging and wearable data collection in days) | 26 (-2,733 ± 319) | 121 (-324 ± 210) | 86 (-506 ± 326) |
| CSF α-synuclein SAA | n participants (Interval between CSF collection and wearable data collection in days) | 26 (-2,702 ± 328) | 114 (-648 ± 463) | 122 (-1,785 ± 979) |

Values are presented as counts or mean ± standard deviation. Intervals represent the time between the DAT scan or CSF collection for SAA testing and the start of wearable data collection period (in days). Negative interval values indicate that the DAT scan or CSF collection was performed before the start of wearable data collection.

### A sleep and activity metrics-based PD score

We computed 32 sleep and activity metrics from raw acceleration signals resampled to 25 Hz (Supplementary Table 2). Temporally aggregating these metrics over observation windows using five statistical measures resulted in 160 candidate features. We then used the computed feature sets from the Verily Study Watch PwPD (n = 133) and HC (n = 26) groups to develop a machine-learning model that distinguishes PwPD from HC. We refer to this model as the SAM-PD model and to its output probability as the SAM-PD score. We first trained, calibrated, and internally evaluated the model under a nested Leave-One-Subject-Out (LOSO) cross-validation scheme. Feature selection within the outer training set of each LOSO fold, based on temporal reliability and collinearity, reduced the feature set to 15 aggregated sleep and activity metrics (‘Machine learning’, Methods). These were combined with age, sex, weight, and height for subsequent model fitting. Of the 160 candidate features, 104 exhibited good test-retest reliability between each participant’s first qualifying fortnight and the subsequent fortnight (ICC(2,1) 95% confidence interval lower bound ≥ 0.75) across folds. Clustering highly correlated features (Spearman’s correlation coefficient |ρ| ≥ 0.90) and retaining the most temporally reliable feature from each cluster produced the final set of 15 features per fold. The consensus feature set across all LOSO folds comprised four sleep, four upper-limb movements and seven ambulatory activity metrics, providing a compact representation of wearable-based sleep and activity digital phenotypes.

Among the classification algorithms evaluated, balanced random forest and ridge logistic regression achieved identical area under the ROC curve (AUROC) (Supplementary Table 3). The former was however selected for SAM-PD because of its lower expected calibration error (ECE). All results reported hereafter correspond to the balanced random forest-based model. Table 3 summarizes the performance of the LOSO-evaluated calibrated and uncalibrated SAM-PD models, with sensitivity and specificity reported at the Youden-optimal operating point [21]. The calibrated model achieved AUROC of 0.93 (95% CI: 0.88–0.97) and an AUPRC of 0.98 (95% CI: 0.97–1.00). Probability calibration reduced the ECE from 0.17 (95% CI: 0.14–0.22) to 0.04 (95% CI: 0.03–0.09). Detailed performance metrics are provided in Supplementary Table 4 and receiver operating characteristic (ROC) and precision-recall curves are shown in Supplementary Fig. 1. In Supplementary Table 4, we also report performance metrics at an operating point that maximizes the F1 score and at an operating point that maximizes sensitivity while maintaining specificity ≥ 0.50, representing a plausible configuration for a screening system. Calibration curves are shown in Supplementary Fig. 2. Hereafter, SAM-PD scores correspond to the calibrated model output probabilities.

**Table 3.** Performance of the uncalibrated and calibrated SAM-PD models in classifying PwPD (n = 133) and HCs (n = 26) in the Verily Study Watch cohort.

| **SAM-PD model** | **AUROC (95% CI)** | **AUPRC (95% CI)** | **Sensitivity (95% CI)** | **Specificity (95% CI)** | **ECE (95% CI)** |
|---|---|---|---|---|---|
| Uncalibrated balanced random forest | 0.92 (0.86–0.97) | 0.98 (0.97–0.99) | 0.86 (0.79–0.92) | 0.81 (0.61–0.93) | 0.17 (0.14–0.22) |
| Calibrated balanced random forest | 0.93 (0.88–0.97) | 0.98 (0.97–1.00) | 0.89 (0.83–0.94) | 0.85 (0.65–0.96) | 0.04 (0.03–0.09) |

Performance is reported using the area under the receiver operating characteristic curve (AUROC), the area under the precision-recall curve (AUPRC), sensitivity, specificity, and expected calibration error (ECE). Positive class (PwPD) prevalence was 83.6%. All metrics are reported as point estimates with 95% confidence intervals (CIs). CIs for sensitivity and specificity are exact Clopper-Pearson binomial intervals, whereas those for all other metrics were obtained from 1,000 bootstrap iterations with replacement. Sensitivity and specificity are reported at the Youden-optimal operating point. Each participant was classified using the decision threshold derived from the out-of-fold predictions of the remaining participants (per-fold leave-one-out approach). Thresholds ranged from 0.46 to 0.49 for the uncalibrated model and from 0.78 to 0.79 for the calibrated model. ECE measures the agreement between predicted probabilities and observed outcome frequencies across probability bins. Lower ECE values indicate better model calibration.

SHapley Additive exPlanations (SHAP)-based feature contributions to the SAM-PD model are shown in Fig. 2. Estimated REM sleep duration was the strongest individual driver of the PD probability, followed by short-term acceleration entropy and peak acceleration. Walking cadence, the variability of short-term acceleration entropy, and estimated REM sleep latency were also among the most predictive features. Among the demographic variables, younger age was associated with a higher PD probability, whereas sex, weight, and height contributed negligibly. This counter-intuitive age association may reflect conditional relationships among predictors or cohort composition and is discussed further in the Discussion section.

**Fig. 2.**
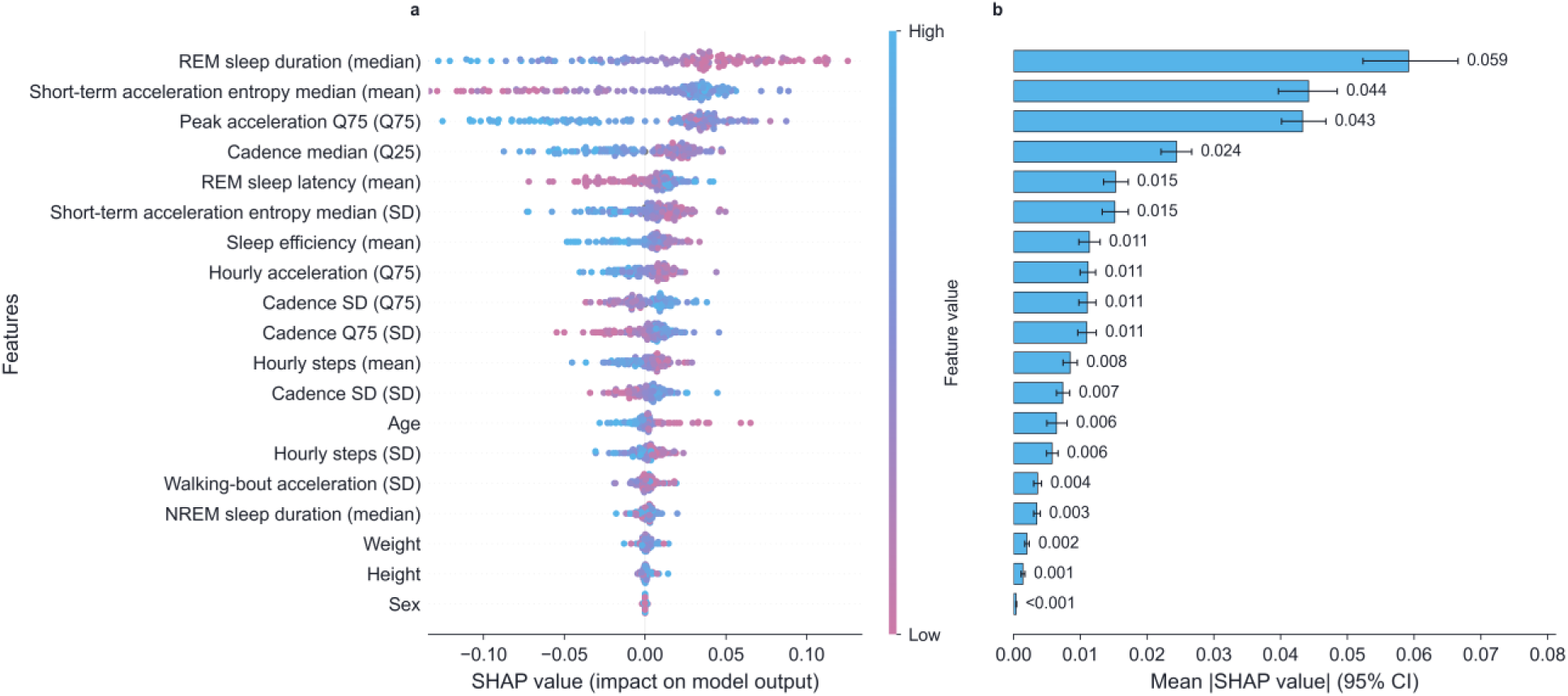
Feature contributions to the output of the SAM-PD model trained on data from PwPD and HCs in the Verily Study Watch cohort. (a) SHAP-based feature contributions computed on held-out test participants during LOSO cross-validation. Each row corresponds to a model input feature and each point to a participant, with color indicating the feature value (high to low). The position on the horizontal axis is determined by the SHAP value, indicating the direction and magnitude of each feature’s contribution to the predicted probability of PD (SAM-PD score); negative SHAP values decrease the SAM-PD score. (b) Feature importance, calculated as the mean absolute SHAP value, with 95% confidence intervals from 1,000 bootstrap resamples with replacement. Features are ordered by their mean absolute SHAP value in both panels. The sleep and activity metrics labels include, in parentheses, the statistical measure used to aggregate the metric over the observation window. For sex, female and male were encoded as 0 and 1, respectively.

We further performed an exploratory assessment of model fairness for the PwPD versus HC classification task by evaluating performance across three attributes, age, sex, and BMI, and calculating disparity metrics, including the Demographic Parity Difference (DPD), Demographic Parity Ratio (DPR), and Equalized Odds Difference (EOD) (Supplementary Tables 5 and 6). Classification performance metrics were computed only for subgroups with at least five participants in each outcome class. AUROC point estimates remained high across evaluable age ranges (50-69 years: AUROC = 0.96, AUPRC = 0.99; ≥ 70 years: AUROC = 0.92, AUPRC = 0.98), as well as between females (AUROC = 0.96) and males (AUROC = 0.90). For BMI subgroups, performance was higher in the overweight subgroup (AUROC = 0.98, AUPRC = 1.00) compared to the normal weight subgroup (AUROC = 0.85, AUPRC = 0.97). Regarding disparity metrics, DPR ranged from 0.86 (age) to 1.00 (BMI), DPD from 0.00 (BMI) to 0.12 (age), and EOD from 0.13 (sex) to 0.33 (BMI). The small number of HCs in certain subgroups limits the precision of these estimates. The cohort was predominantly White (n = 153) and of unspecified ethnicity (n = 110), whereas all other racial and ethnic subgroups were underrepresented, precluding meaningful fairness evaluation for these attributes.

Performance is reported using the area under the receiver operating characteristic curve (AUROC), the area under the precision-recall curve (AUPRC), sensitivity, specificity, and expected calibration error (ECE). Positive class (PwPD) prevalence was 83.6%. All metrics are reported as point estimates with 95% confidence intervals (CIs). CIs for sensitivity and specificity are exact Clopper-Pearson binomial intervals, whereas those for all other metrics were obtained from 1,000 bootstrap iterations with replacement. Sensitivity and specificity are reported at the Youden-optimal operating point. Each participant was classified using the decision threshold derived from the out-of-fold predictions of the remaining participants (per-fold leave-one-out approach). Thresholds ranged from 0.46 to 0.49 for the uncalibrated model and from 0.78 to 0.79 for the calibrated model. ECE measures the agreement between predicted probabilities and observed outcome frequencies across probability bins. Lower ECE values indicate better model calibration.

### Association with dopaminergic deficit and α-synuclein pathology

Having established that SAM-PD effectively discriminates PwPD from HCs, we next assessed whether the model captures the presence of dopaminergic deficit and α-synuclein pathology in the Verily Study Watch at-risk group. To this end, we retrained and calibrated the SAM-PD model using the complete Verily Study Watch PwPD (n = 133) and HC (n = 26) dataset, with the consensus feature set and hyperparameters derived from the LOSO cross-validation, and then applied it to the at-risk individuals without any additional tuning. We further compared the performance of SAM-PD with two versions of the MDS prodromal PD risk score: a version incorporating all available risk and prodromal markers, and a mini version incorporating only markers that do not require specialized testing (see Methods, ‘MDS prodromal PD risk score and combination strategies’).

Figure 3 illustrates the relationship between biomarkers of dopaminergic deficit (DAT scan-derived minimum putaminal specific binding ratio [SBR]) and α-synuclein pathology (CSF α-synuclein SAA mean maximum fluorescence [Fmax]) and the SAM-PD and MDS scores. In the combined PwPD and HC cohort (Fig. 3a), higher SAM-PD scores were associated with lower SBR (Pearson r = −0.60, p < 0.05, n = 112) and higher SAA mean Fmax (r = +0.48, p < 0.05, n = 128); 20 of 148 participants were excluded from the Fmax correlation analysis, because their SAA results were obtained using a different assay protocol. In the at-risk group (Fig. 3b), the SAM-PD score showed weak-to-moderate, statistically significant correlations with both biomarkers (SBR: r = −0.44, p < 0.05, n = 121; mean Fmax: r = +0.40, p < 0.05, n = 98) in the expected direction. The MDS score showed similarly significant correlations (SBR: r = −0.33, p < 0.05, n = 121; mean Fmax: r = +0.31, p < 0.05, n = 98), whereas the mini-MDS risk score correlated significantly, albeit weakly, only with SBR (r = −0.18, p < 0.05, n = 121).

**Fig. 3.**
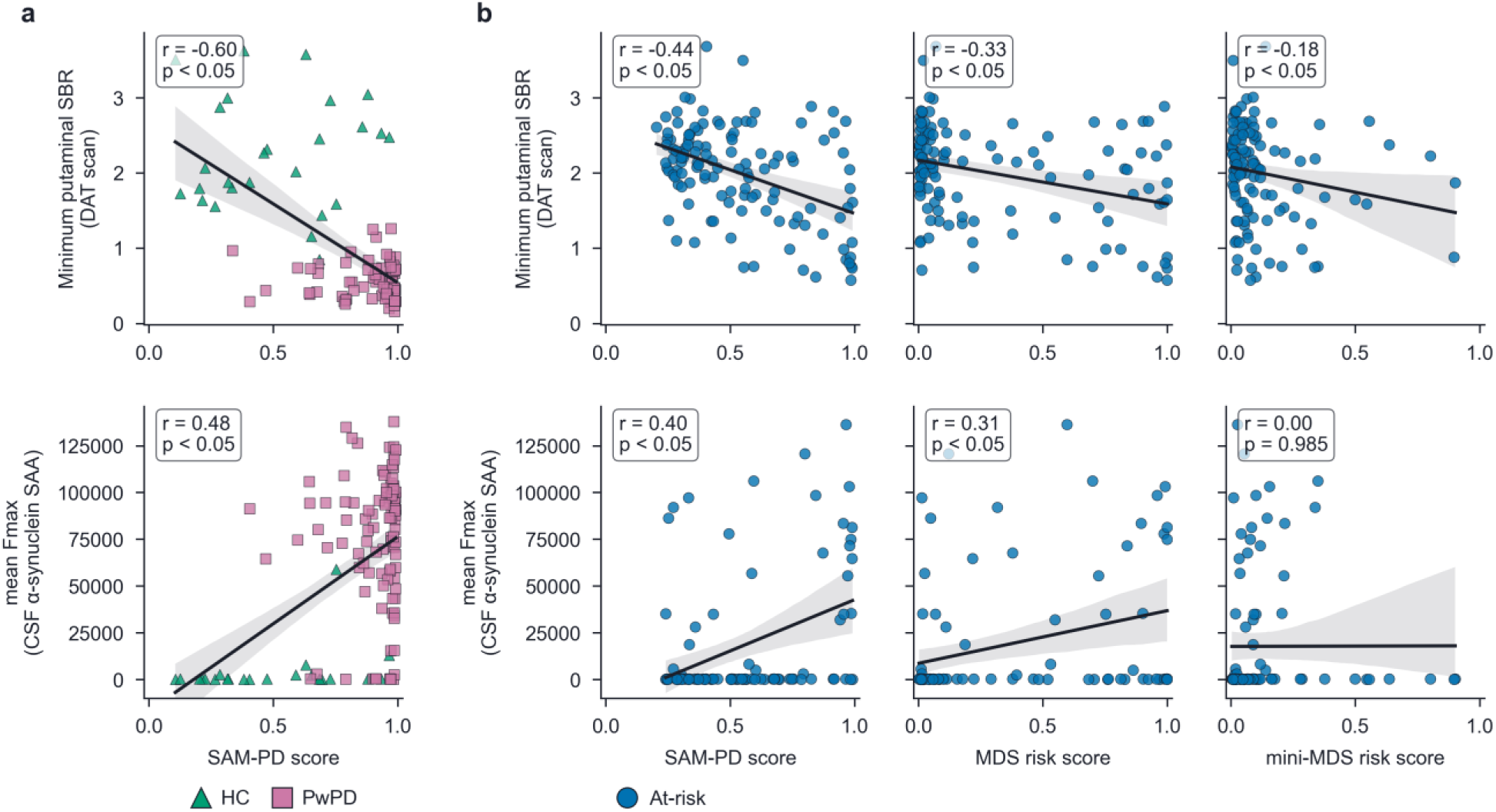
Relationship between the SAM-PD and MDS prodromal PD risk scores and biomarkers of dopaminergic deficit and α-synuclein pathology in the Verily Study Watch cohort. (a) Scatter plots of the DAT scan-derived minimum putaminal SBR (top) and the CSF α-synuclein SAA mean Fmax (bottom) against the SAM-PD score in the combined PwPD and HC groups. (b) Scatter plots of the DAT scan-derived minimum putaminal SBR (top) and the CSF α-synuclein SAA mean Fmax (bottom) against the SAM-PD score, the MDS risk score, and the mini-MDS risk score (columns, left to right) in the at-risk group. Mean Fmax is the mean maximum fluorescence across three SAA replicates. In each panel, the least-squares regression line and its 95% confidence interval (shaded area) are shown; the inset displays the Pearson correlation coefficient (r) and corresponding p value.

Table 4 summarizes the performance of the SAM-PD and MDS scores in detecting the presence of dopaminergic deficit and α-synuclein pathology in the at-risk group. ROC and precision-recall curves are shown in Supplementary Figs. 3 and 4, respectively, and additional performance metrics are included in Supplementary Table 7. For dopaminergic deficit, the SAM-PD score showed higher AUROC but comparable AUPRC to the MDS risk score in distinguishing individuals with normal versus abnormal DAT scans (AUROC = 0.86, 95% CI: 0.75–0.94; AUPRC = 0.45, 95% CI: 0.25–0.73 vs. AUROC = 0.80, 95% CI: 0.66–0.91; AUPRC = 0.46, 95% CI: 0.21–0.68). For α-synuclein pathology, the SAM-PD model showed comparable AUROC and higher AUPRC than the MDS risk score in distinguishing individuals with negative versus positive SAA (AUROC = 0.76, 95% CI: 0.62–0.90; AUPRC = 0.50, 95% CI: 0.30– 0.75 vs. AUROC = 0.76, 95% CI: 0.65–0.87; AUPRC = 0.42, 95% CI: 0.25–0.63). SAM-PD detected the presence of at least one pathological feature with an AUROC of 0.79 (95% CI: 0.67–0.90) and AUPRC of 0.59 (95% CI: 0.40–0.79), demonstrating higher AUPRC and comparable AUROC to the MDS risk score (AUROC = 0.76, 95% CI: 0.65–0.85; AUPRC = 0.50, 95% CI: 0.35–0.68). In identifying individuals with both pathological features, SAM-PD achieved an AUROC of 0.94 (95% CI: 0.86–1.00) and an AUPRC of 0.57 (95% CI: 0.30–0.96), performing comparably to the MDS risk score (AUROC = 0.93, 95% CI: 0.86– 0.98; AUPRC = 0.56, 95% CI: 0.23–0.85). Across all four pathological hallmark scenarios, AUROC and AUPRC were consistently higher for SAM-PD than the mini-MDS risk score.

**Fig. 4.**
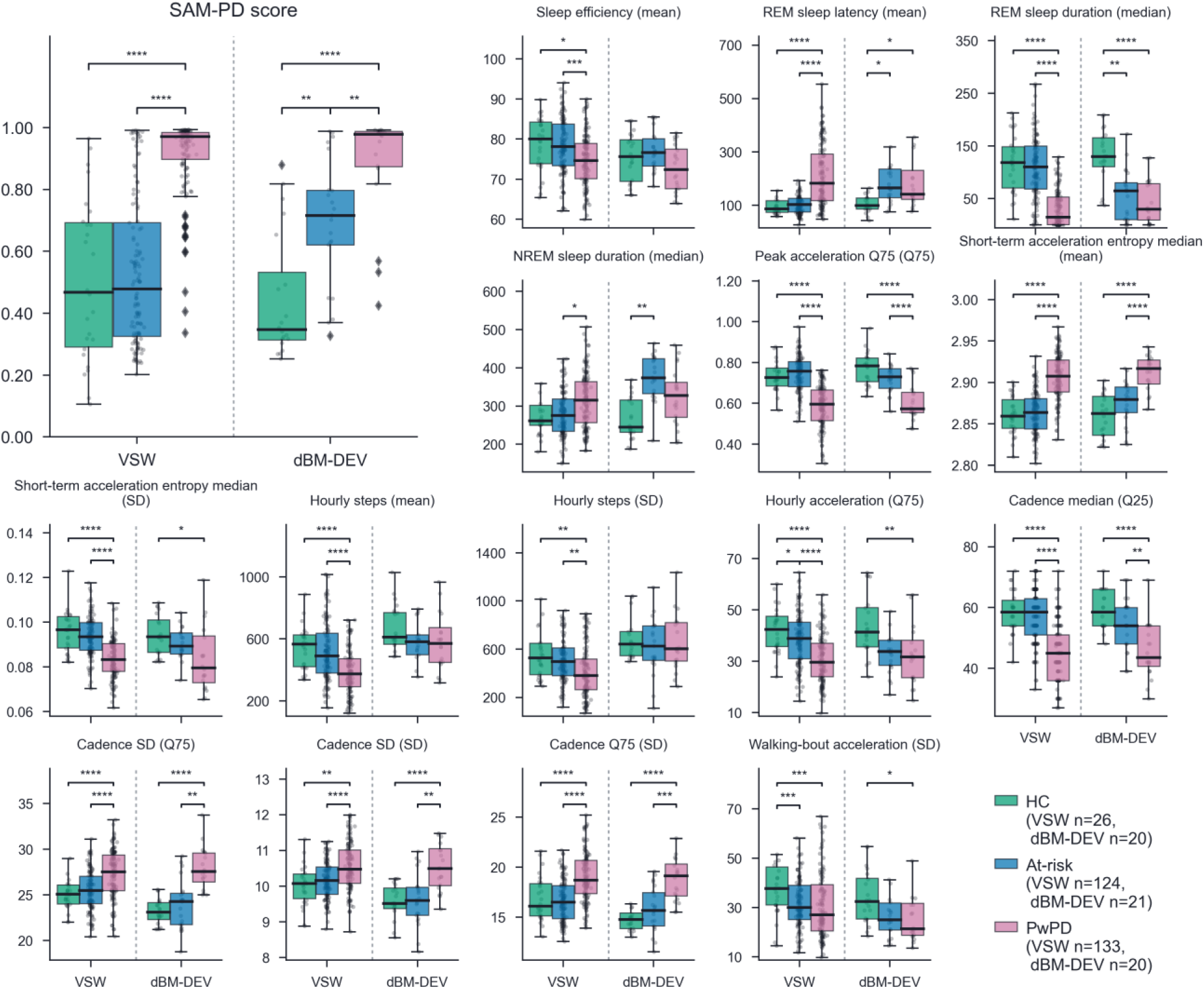
SAM-PD score and input feature distributions in the Verily Study Watch and dBM-DEV cohorts. Box plots show the distribution of the SAM-PD score (large panel) and each of its 15 constituent sleep and activity metrics in the HC, at-risk, and PwPD groups of both cohorts [Verily Study Watch (VSW): HC n = 26, at-risk n = 124, PwPD n = 133; dBM-DEV: HC n = 20, at-risk n = 21, PwPD n = 20], separated by the vertical dashed line. Boxes denote the interquartile range (IQR) with the median indicated by the horizontal line within each box; whiskers extend to 1.5 × IQR, and overlaid grey points represent individual participant values. Statistically significant pairwise differences are annotated (*, p < 0.05; **, p < 0.01; ***, p < 0.001; ****, p < 0.0001). Statistical comparisons were performed using rank-based ANCOVA for sleep and activity metrics, with age and sex adjustment, and the two-sided Mann-Whitney U test for the SAM-PD score. All p values were Bonferroni-corrected. The SAM-PD model was applied to the independent dBM-DEV cohort without retraining or recalibration.

**Table 4.**
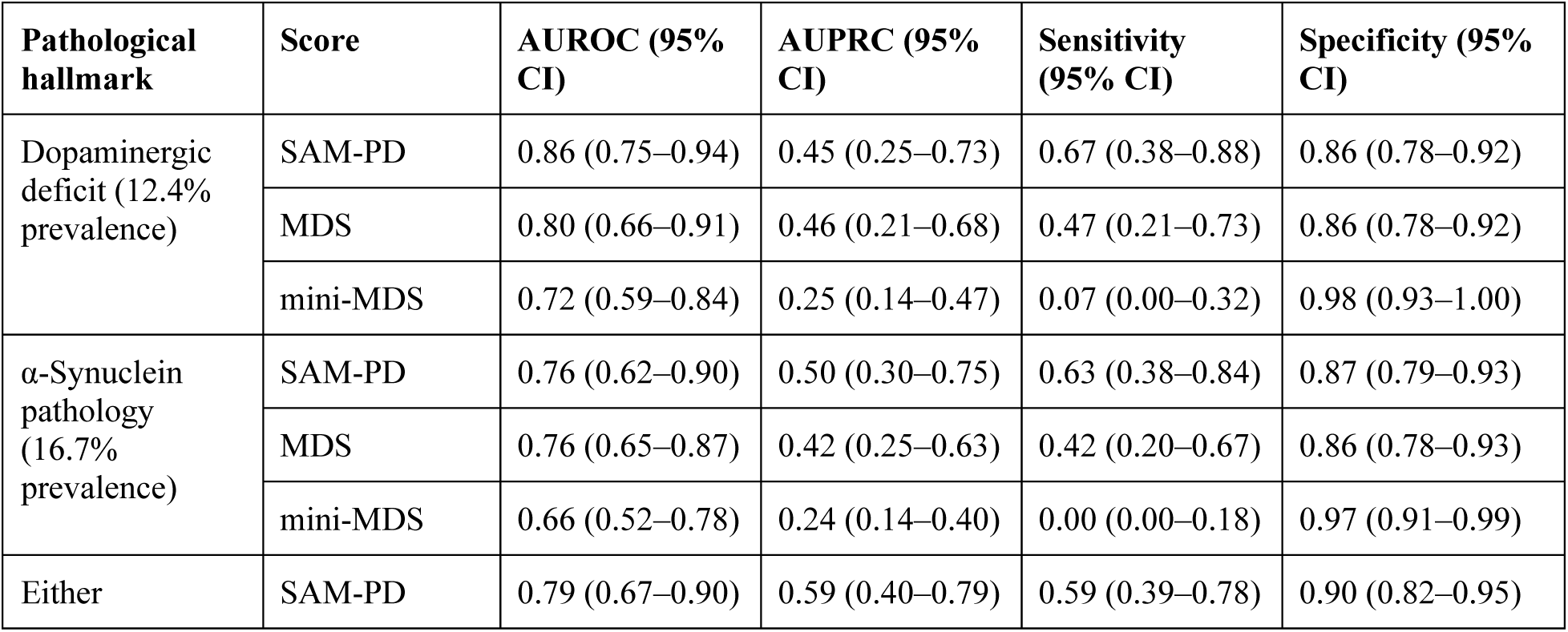
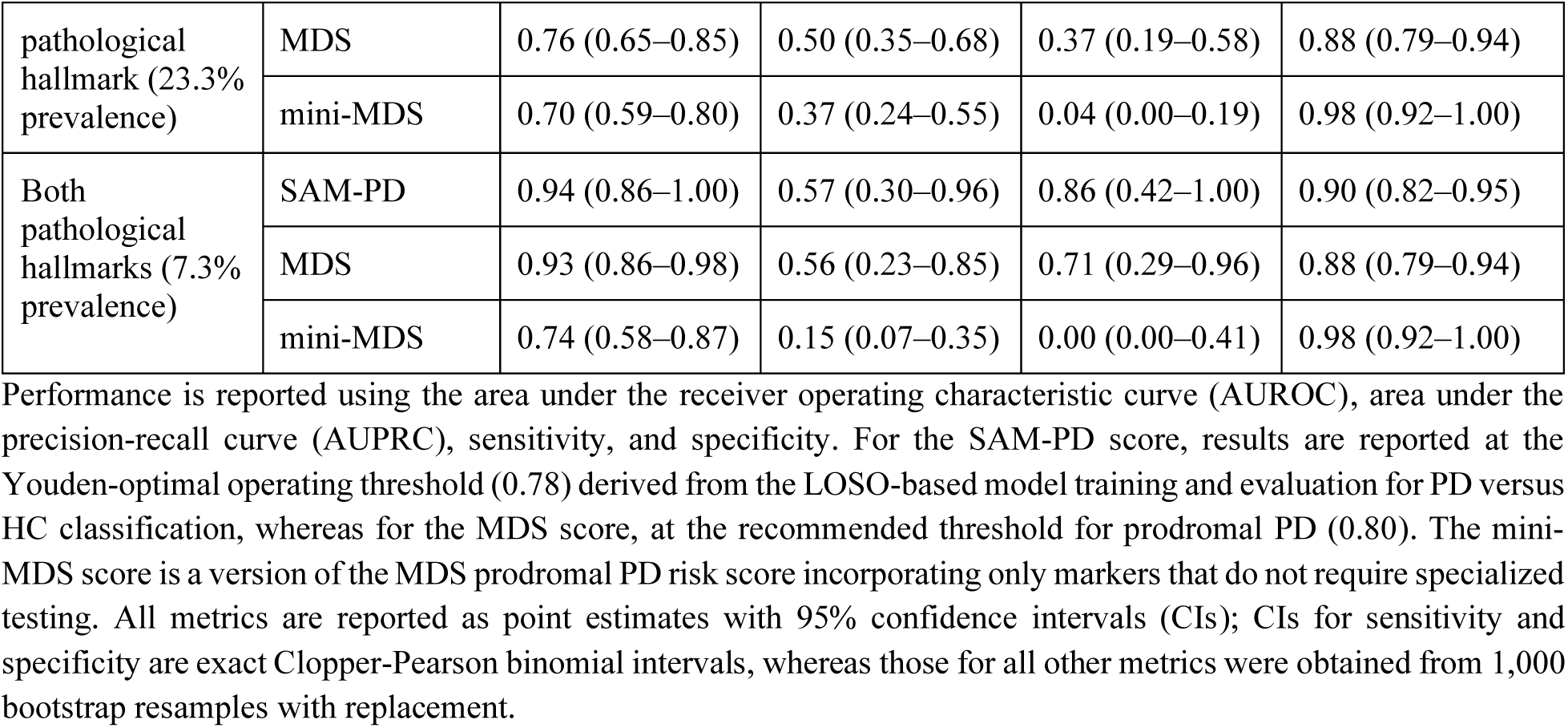
Performance of the SAM-PD and MDS prodromal PD risk scores for detecting pathological hallmarks in the Verily Study Watch at-risk group.

| **Pathological hallmark** | **Score** | **AUROC (95% CI)** | **AUPRC (95% CI)** | **Sensitivity (95% CI)** | **Specificity (95% CI)** |
|---|---|---|---|---|---|
| Dopaminergic deficit (12.4% prevalence) | SAM-PD | 0.86 (0.75–0.94) | 0.45 (0.25–0.73) | 0.67 (0.38–0.88) | 0.86 (0.78–0.92) |
|  | MDS | 0.80 (0.66–0.91) | 0.46 (0.21–0.68) | 0.47 (0.21–0.73) | 0.86 (0.78–0.92) |
|  | mini-MDS | 0.72 (0.59–0.84) | 0.25 (0.14–0.47) | 0.07 (0.00–0.32) | 0.98 (0.93–1.00) |
| α-Synuclein pathology (16.7% prevalence) | SAM-PD | 0.76 (0.62–0.90) | 0.50 (0.30–0.75) | 0.63 (0.38–0.84) | 0.87 (0.79–0.93) |
|  | MDS | 0.76 (0.65–0.87) | 0.42 (0.25–0.63) | 0.42 (0.20–0.67) | 0.86 (0.78–0.93) |
|  | mini-MDS | 0.66 (0.52–0.78) | 0.24 (0.14–0.40) | 0.00 (0.00–0.18) | 0.97 (0.91–0.99) |
| Either pathological hallmark (23.3% prevalence) | SAM-PD | 0.79 (0.67–0.90) | 0.59 (0.40–0.79) | 0.59 (0.39–0.78) | 0.90 (0.82–0.95) |
|  | MDS | 0.76 (0.65–0.85) | 0.50 (0.35–0.68) | 0.37 (0.19–0.58) | 0.88 (0.79–0.94) |
|  | mini-MDS | 0.70 (0.59–0.80) | 0.37 (0.24–0.55) | 0.04 (0.00–0.19) | 0.98 (0.92–1.00) |
| Both pathological hallmarks (7.3% prevalence) | SAM-PD | 0.94 (0.86–1.00) | 0.57 (0.30–0.96) | 0.86 (0.42–1.00) | 0.90 (0.82–0.95) |
|  | MDS | 0.93 (0.86–0.98) | 0.56 (0.23–0.85) | 0.71 (0.29–0.96) | 0.88 (0.79–0.94) |
|  | mini-MDS | 0.74 (0.58–0.87) | 0.15 (0.07–0.35) | 0.00 (0.00–0.41) | 0.98 (0.92–1.00) |

Performance is reported using the area under the receiver operating characteristic curve (AUROC), area under the precision-recall curve (AUPRC), sensitivity, and specificity. For the SAM-PD score, results are reported at the Youden-optimal operating threshold (0.78) derived from the LOSO-based model training and evaluation for PD versus HC classification, whereas for the MDS score, at the recommended threshold for prodromal PD (0.80). The mini-MDS score is a version of the MDS prodromal PD risk score incorporating only markers that do not require specialized testing. All metrics are reported as point estimates with 95% confidence intervals (CIs); CIs for sensitivity and specificity are exact Clopper-Pearson binomial intervals, whereas those for all other metrics were obtained from 1,000 bootstrap resamples with replacement.

For the SAM-PD score, the decision threshold was set at the Youden-optimal operating point (0.78) derived from the LOSO-based model training and evaluation for PwPD versus HC classification. For the MDS score, the recommended threshold for prodromal PD (0.80) was used [8]. SAM-PD demonstrated the highest sensitivity across all pathological hallmark scenarios while maintaining a specificity consistently ≥ 0.85, comparable to that of the MDS score. The mini-MDS score yielded the highest specificity across all four scenarios, but had very low sensitivity, indicating limited case detection at the recommended 0.80 threshold. Supplementary Fig. 5 shows the distributions of the SAM-PD, MDS, and mini-MDS scores across pathological hallmark scenarios in Verily Study Watch at-risk individuals. Pairwise comparisons using a two-sided Mann-Whitney U test yielded statistically significant differences between the positive and negative groups across all scenarios for all three scores.

Performance is reported using the area under the receiver operating characteristic curve (AUROC), area under the precision-recall curve (AUPRC), sensitivity, and specificity. For the SAM-PD score, results are reported at the Youden-optimal operating threshold (0.78) derived from the LOSO-based model training and evaluation for PD versus HC classification, whereas for the MDS score, at the recommended threshold for prodromal PD (0.80). The mini-MDS score is a version of the MDS prodromal PD risk score incorporating only markers that do not require specialized testing. All metrics are reported as point estimates with 95% confidence intervals (CIs); CIs for sensitivity and specificity are exact Clopper-Pearson binomial intervals, whereas those for all other metrics were obtained from 1,000 bootstrap resamples with replacement.

### External validation in independent cohort

To assess its generalizability, we applied the SAM-PD model, trained using the PwPD and HC cohorts of the Verily Study Watch dataset, to the independent dBM-DEV cohort (n = 61) without additional tuning. The same decision threshold (0.78) as in the at-risk analyses was used. Despite differences in hardware and geographic location, SAM-PD demonstrated similar performance in distinguishing PwPD (n = 20) from HCs (n = 20), achieving an AUROC of 0.95 (95% CI: 0.87–0.99) and an AUPRC of 0.95 (95% CI: 0.89–0.99), with a sensitivity of 0.85 (95% CI: 0.62–0.97) and a specificity of 0.90 (95% CI: 0.68-0.99). ROC and precision-recall curves are shown in Supplementary Fig. 6, whereas detailed performance metrics, including those at alternative decision thresholds, are provided in Supplementary Table 8.

Figure 4 shows the distributions of the SAM-PD score and its constituent sleep and activity metrics across the PwPD, at-risk, and HC groups in the Verily Study Watch and dBM-DEV cohorts. Statistical comparisons were performed using rank-based analysis of covariance (ANCOVA) for sleep and activity metrics, with age and sex adjustment, and the two-sided Mann-Whitney U test for the SAM-PD score. In both cases, Bonferroni correction was applied. In the Verily Study Watch cohort, the SAM-PD score and input feature values of at-risk individuals generally fell between those of PwPD and HCs. Pairwise comparisons revealed statistically significant differences between the PwPD group and each of the other two groups for all measures except non-REM (NREM) sleep duration and walking-bout acceleration. No significant differences were observed between HCs and at-risk individuals, except for walking-bout acceleration and hourly acceleration. Similar patterns were observed for the SAM-PD score and most input features in the dBM-DEV cohort. Nevertheless, for nearly half of the metrics, the differences between PwPD and each of the other two groups were not statistically significant. Notably, in this cohort, at-risk individuals, all of whom had PSG-confirmed RBD, also exhibited significantly shorter REM sleep duration, longer REM sleep latency, and longer NREM sleep duration than HCs, whereas these sleep metrics were comparable to those of PwPD. The detailed adjusted and unadjusted comparisons are provided in Supplementary Table 9.

### Test-retest reliability

We next evaluated the temporal reliability of the SAM-PD score and its constituent sleep and activity metrics by conducting a test-retest analysis in both the Verily Study Watch (n = 119) and dBM-DEV (n = 50) cohorts. To characterize how reliability changes with longer monitoring, we computed the absolute-agreement, single-measurement intraclass correlation coefficient (ICC(2,1)) over consecutive, non-overlapping windows of 3, 7, 10, and 14 days spanning each participant’s first qualifying fortnight and the immediately subsequent fortnight, assuming that participants’ health status remained stable throughout this interval. Figure 5 shows the evolution of ICC(2,1) across observation windows for the SAM-PD score and its input features in the Verily Study Watch and dBM-DEV cohorts. In both cohorts, reliability improved with longer aggregation windows, with sleep and hourly step features showing the greatest improvement. The selected Verily Study Watch features were required to meet the good-to-excellent reliability criterion (ICC(2,1) 95% CI lower bound ≥ 0.75) at the 14-day window, and all also showed good-to-excellent reliability in the dBM-DEV cohort (ICC(2,1) ≥ 0.75). The SAM-PD score achieved excellent absolute agreement in both cohorts when computed from features aggregated over 14 days (Verily Study Watch: ICC(2,1) = 0.96, 95% CI: 0.94–0.97; dBM-DEV: ICC(2,1) = 0.96, 95% CI: 0.93–0.98). Supplementary Table 10 reports ICC(2,1) values for all input features and the SAM-PD score in both cohorts, together with the relative importance of each feature in the model.

**Fig. 5.**
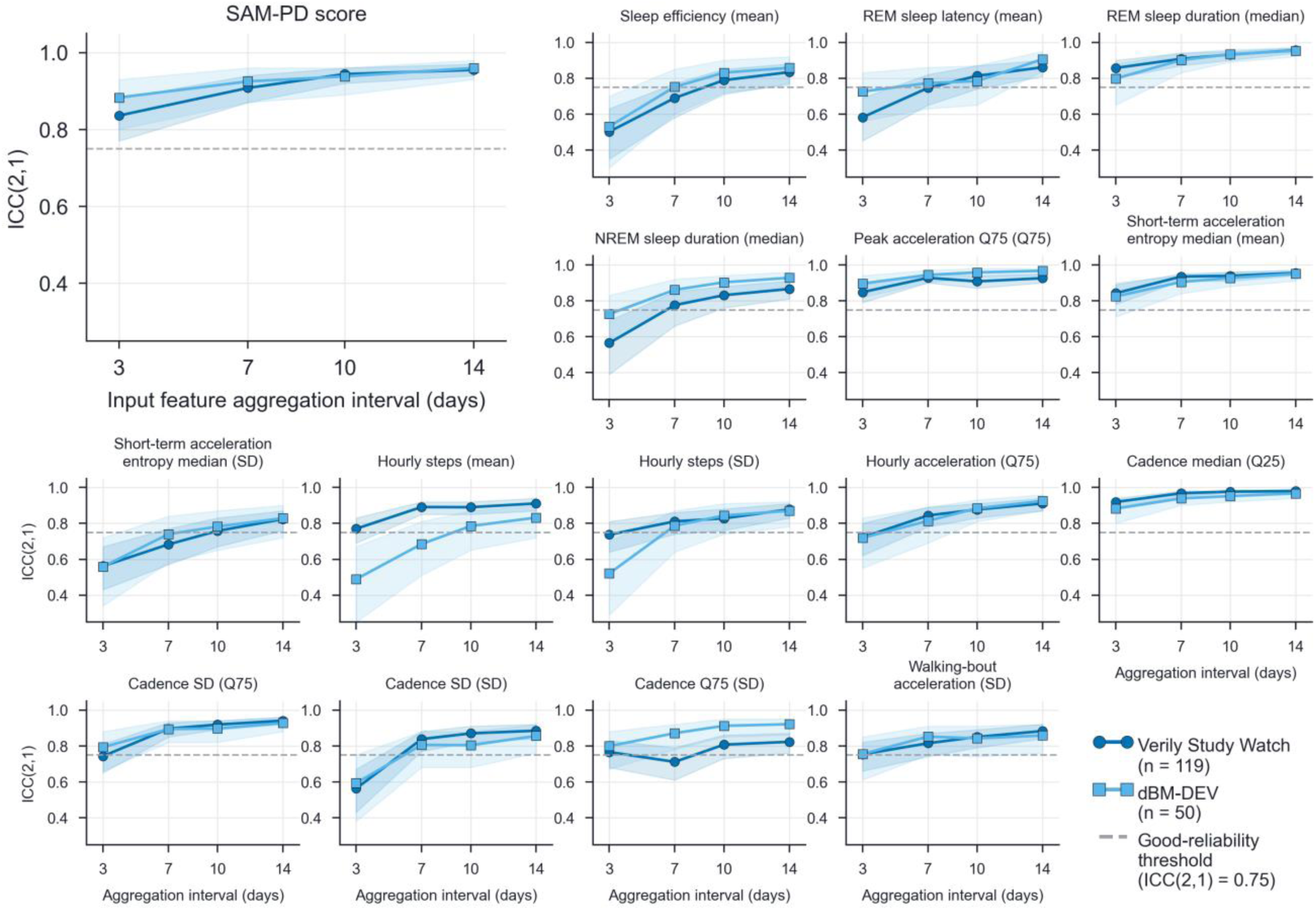
Test-retest reliability of the SAM-PD score and its constituent sleep and activity metrics across feature aggregation intervals in the Verily Study Watch and dBM-DEV cohorts. The absolute-agreement, single-measurement intraclass correlation coefficient ICC(2,1) is plotted against the feature aggregation interval (3, 7, 10, and 14 days) for the SAM-PD score (large panel) and each of its 15 constituent sleep and activity metrics, in the Verily Study Watch (n = 119) and dBM-DEV (n = 50) cohorts. Shaded areas indicate 95% confidence intervals, and the horizontal dashed line marks the good-reliability threshold (ICC(2,1) = 0.75).

### Complementarity with the MDS prodromal PD risk score

To explore the complementarity of the SAM-PD and MDS scores, we first examined their correlation in the at-risk group of the Verily Study Watch cohort. The SAM-PD score showed a moderate positive correlation with the MDS risk score (r = 0.43, p < 0.05) and a weak, non-significant positive correlation with the mini-MDS risk score (r = 0.17, p = 0.06). Together with their comparative performance in detecting pathological features, these findings suggest that the SAM-PD and MDS scores may capture complementary aspects of individuals’ health status.

We then investigated whether combining the SAM-PD and the MDS scores could improve the detection of pathological hallmarks. We evaluated several score-combination strategies (‘MDS prodromal PD risk score and combination strategies’, Methods), including fusion into an Ensemble PD score using a meta-learner, maximum-score screening, and sequential screening. Table 5 summarizes the performance for the Ensemble PD score, which consistently yielded higher performance metrics than the other two approaches, whereas performance metrics for all combination strategies are reported in Supplementary Table 7. ROC and precision-recall curves are shown in Supplementary Figs. 3 and 4, respectively. For dopaminergic deficit, the Ensemble PD score achieved an AUROC of 0.89 (95% CI: 0.80–0.96) and an AUPRC of 0.61 (95% CI: 0.35–0.83), whereas for α-synuclein pathology, it achieved an AUROC of 0.77 (95% CI: 0.64–0.91) and an AUPRC of 0.56 (95% CI: 0.36–0.79). The Ensemble PD score also achieved an AUROC of 0.81 (95% CI: 0.70–0.91; AUPRC = 0.67, 95% CI: 0.50–0.83) for detecting the presence of either pathological hallmark and an AUROC of 0.97 (95% CI: 0.90–1.00; AUPRC = 0.81, 95% CI: 0.50– 1.00) for detecting the concurrent presence of both disease hallmarks. Compared with the standalone scores (Table 4), the Ensemble PD score had a higher or equal AUROC in all four pathological hallmark scenarios and the highest AUPRC in each scenario; however, the corresponding 95% confidence intervals overlapped with those of the standalone scores. The mini-Ensemble PD score, obtained by fusing the SAM-PD and mini-MDS scores, achieved AUROC estimates comparable to those of SAM-PD alone and at least as high as those of both MDS score versions, yet consistently lower AUPRC estimates than the Ensemble PD score. The fitted meta-learner coefficients are reported in Supplementary Table 11. In both meta-learners, the SAM-PD score received greater weight than the MDS or mini-MDS risk scores.

**Table 5.** Performance of the meta-learner-based Ensemble PD score, fusing the SAM-PD and MDS scores, for detecting pathological hallmarks in the Verily Study Watch at-risk group.

| **Pathological hallmark** | **Score** | **AUROC (95% CI)** | **AUPRC (95% CI)** | **Sensitivity (95% CI)** | **Specificity (95% CI)** |
|---|---|---|---|---|---|
| Dopaminergic deficit (12.4% prevalence) | Ensemble PD score | 0.89 (0.80–0.96) | 0.61 (0.35–0.83) | 0.60 (0.32–0.84) | 0.89 (0.81–0.94) |
|  | Mini-Ensemble PD score | 0.86 (0.76–0.94) | 0.46 (0.23–0.71) | 0.80 (0.52–0.96) | 0.81 (0.72–0.88) |
| α-Synuclein pathology (16.7% prevalence) | Ensemble PD score | 0.77 (0.64–0.91) | 0.56 (0.36–0.79) | 0.58 (0.33–0.80) | 0.89 (0.81–0.95) |
|  | Mini-Ensemble PD score | 0.77 (0.65–0.89) | 0.45 (0.28–0.73) | 0.63 (0.38–0.84) | 0.80 (0.71–0.88) |
| Either pathological hallmark (23.3% prevalence) | Ensemble PD score | 0.81 (0.70–0.91) | 0.67 (0.50–0.83) | 0.52 (0.32–0.71) | 0.92 (0.84–0.97) |
|  | Mini-Ensemble PD score | 0.80 (0.69–0.90) | 0.60 (0.41–0.79) | 0.67 (0.46–0.83) | 0.84 (0.75–0.91) |
| Both pathological hallmarks (7.3% prevalence) | Ensemble PD score | 0.97 (0.90–1.00) | 0.81 (0.50–1.00) | 0.86 (0.42–1.00) | 0.92 (0.84–0.97) |
|  | Mini-Ensemble PD score | 0.94 (0.88–0.99) | 0.53 (0.28–0.92) | 0.86 (0.42–1.00) | 0.84 (0.75–0.91) |

Performance is reported using the area under the receiver operating characteristic curve (AUROC), area under the precision-recall curve (AUPRC), sensitivity, and specificity, each at the Youden-optimal operating threshold of the corresponding meta-learner (0.60 for the Ensemble PD score and 0.43 for the mini-Ensemble PD score). The mini-Ensemble PD score is the fusion of the SAM-PD and mini-MDS scores. All metrics are reported as point estimates with 95% confidence intervals (CIs); CIs for sensitivity and specificity are exact Clopper-Pearson binomial intervals, whereas those for all other metrics were obtained from 1,000 bootstrap resamples with replacement.

The Ensemble PD score achieved higher sensitivity than the standalone MDS risk score across all pathological hallmark scenarios (0.52–0.86 vs. 0.37–0.71), while maintaining comparable specificity (0.89–0.92 vs. 0.86–0.88). For detecting the concurrent presence of both disease hallmarks, both ensemble scores reached a sensitivity of 0.86; however, this result should be interpreted cautiously given the small number of positive cases (n = 7). The mini-Ensemble PD score matched or exceeded the Ensemble PD score in sensitivity in all four scenarios, while showing lower specificity. In Supplementary Table 12, we also report performance metrics at an operating point that maximizes the F1 score and at an operating point that maximizes sensitivity while maintaining specificity ≥ 0.50. Supplementary Fig. 5 shows the distributions of both ensemble scores across pathological hallmark scenarios in Verily Study Watch at-risk individuals. Pairwise comparisons using a two-sided Mann-Whitney U test yielded statistically significant differences between the positive and negative groups across all scenarios for both scores.

Supplementary Fig. 7 shows the relationship between the biomarkers of dopaminergic deficit (DAT scan-derived minimum putaminal SBR) and α-synuclein pathology (CSF α-synuclein SAA mean Fmax) and the Ensemble and mini-Ensemble PD scores. Both scores showed weak-to-moderate, statistically significant correlations with both biomarkers in the expected directions, with the Ensemble PD score demonstrating slightly higher absolute correlation coefficients (SBR: r = −0.47, p < 0.05, n = 121; mean Fmax: r = +0.47, p < 0.05, n = 98) than the mini-Ensemble PD score (SBR: r = −0.46, p < 0.05, n = 121; mean Fmax: r = +0.40, p < 0.05, n = 98).

Performance is reported using the area under the receiver operating characteristic curve (AUROC), area under the precision-recall curve (AUPRC), sensitivity, and specificity, each at the Youden-optimal operating threshold of the corresponding meta-learner (0.60 for the Ensemble PD score and 0.43 for the mini-Ensemble PD score). The mini-Ensemble PD score is the fusion of the SAM-PD and mini-MDS scores. All metrics are reported as point estimates with 95% confidence intervals (CIs); CIs for sensitivity and specificity are exact Clopper-Pearson binomial intervals, whereas those for all other metrics were obtained from 1,000 bootstrap resamples with replacement.

## Discussion

This study demonstrates that passive sleep and activity tracking using wrist accelerometry can effectively discriminate PwPD from HCs and identify at-risk individuals with dopaminergic deficit or α-synuclein pathology. Key strengths include the generalizability of the SAM-PD score to an independent cohort, the good-to-excellent temporal reliability of both the score and its constituent sleep and activity metrics, and its complementarity with the MDS prodromal PD risk score. All sleep and activity metrics used in this study were derived from raw accelerometer signals using open-source algorithms, further supporting reproducibility and independent validation across new cohorts and wearable hardware platforms.

We framed model development as a PwPD-versus-HC classification task because our primary aim was to develop a PD screening system rather than models for detecting the specific pathological hallmarks.

We then evaluated the SAM-PD model in at-risk individuals, considering the presence of dopaminergic deficit or α-synuclein pathology as biologically relevant markers of future phenoconversion [22–24]. Importantly, the vast majority of PwPD in the training set had objective evidence of nigrostriatal degeneration and α-synuclein pathology: 98.8% (85/86) exhibited a dopaminergic deficit on DAT imaging and 86.1% (105/122) had a positive SAA test (Fig. 1). In contrast, none of the 26 HCs had an abnormal DAT scan result or a positive SAA test. These findings support the biological relevance of the clinical labels used for model development and motivate evaluation of the SAM-PD score against biomarker status in at-risk individuals.

Sleep and activity metrics were selected to build the core SAM-PD model for distinguishing PwPD from HCs because both domains are affected by the disease and encompass early, subclinical manifestations that may go unnoticed. Indeed, sleep disturbances [25] and subtle upper-limb motor changes [26] are among the earliest signs of the disorder that can be captured unobtrusively using wrist accelerometry. In this study, the most predictive features comprised a combination of sleep, upper-limb movement, and ambulatory metrics that showed biologically plausible contributions to the SAM-PD score. Shorter REM sleep duration was the strongest predictor and, together with reduced sleep efficiency, reduced estimated NREM duration and longer estimated REM sleep latency, increased the score, consistent with sleep disturbances commonly observed in PD. REM sleep time was also identified as a significant predictor in a similar study by Schalkamp et al. (2025) [15], which used precomputed sleep features derived from a different sleep-staging algorithm. Moreover, the contribution of salient upper-limb movement and ambulatory metrics to the SAM-PD output is consistent with the motor impairment associated with PD. Higher short-term acceleration entropy, indicating a more irregular movement intensity during brief hand-movement bouts, increased the SAM-PD score. This may reflect the increased variability of upper-limb movements described in PD, which has been linked to bradykinesia and is most pronounced for fast, brief movements [27]. Features reflecting reduced peak acceleration and lower hourly acceleration, as well as lower variability in walking-bout acceleration, further increased the score. These findings underscore the importance of reduced movement vigor, overall activity volume, and constrained movement patterns, respectively. In addition, reduced and more variable walking cadence, indicative of slower and less rhythmic stepping, together with reduced and less variable hourly step counts, increased the SAM-PD score. This pattern is consistent with impaired gait and decreased, more stereotyped daily activity that is common in PD [28].

While advanced age is an established risk factor for PD [29], our SHAP-based analysis indicated that younger age was associated with an increased predicted probability of PD within the SAM-PD model. This counter-intuitive association may reflect conditional relationships among predictors or cohort composition. The SAM metrics used in our model may vary with age independently of disease status. In this context, demographic variables may serve to contextualize these metrics rather than act as isolated predictors. For instance, older age is naturally associated with reduced activity and altered sleep patterns, regardless of disease status. Therefore, a lower activity profile in a younger individual may represent a greater deviation from the expected age-specific baseline compared with a similar profile in an older individual. Alternatively, this finding may partly reflect the underrepresentation of HCs in younger age subgroups of the development cohort. Further investigation in larger and more age-balanced populations is needed to determine the reproducibility and underlying drivers of this age association.

For detecting PD, the SAM-PD score achieved an AUROC of 0.93 (AUPRC = 0.98) in the U.S.-based Verily Study Watch cohort. Similar high discrimination was also observed in the European dBM-DEV study cohort (AUROC = 0.95; AUPRC = 0.95), which was monitored using a different, consumer-grade smartwatch. The findings support the geographic and cross-device generalizability of SAM-PD.

Performance in the Verily Study Watch cohort was slightly higher than that reported by Schalkamp et al. (2025) [15], who used a logistic regression model trained on sleep, physical activity, and vital sign data (AUPRC = 0.96). However, differences in participant selection and model evaluation between the studies preclude direct performance comparison. The SAM-PD score and input feature values of at-risk individuals generally fell between those of PwPD and HCs. Pairwise comparisons in the Verily Study Watch cohort showed statistically significant differences between the PwPD group and the two other groups in almost all cases. Such differences were largely absent between HCs and at-risk individuals, which is to be expected, as not all at-risk individuals had confirmed neuropathological features of interest or other pathologies that could affect sleep or activity. This was not the case for the dBM-DEV cohort, where all at-risk individuals had PSG-confirmed RBD. Although similar patterns to those of the Verily Study Watch cohort were observed in most upper-limb movement and ambulatory metrics of the three groups, at-risk individuals in the dBM-DEV cohort exhibited significantly shorter estimated REM sleep duration, longer estimated REM sleep latency, and longer estimated NREM sleep duration than HCs, whereas these sleep metrics were comparable to those of PwPD.

Temporal reliability analyses further showed that two weeks of data were sufficient to produce reliable (ICC(2,1) > 0.75) sleep and activity input metrics, as well as SAM-PD scores, in both cohorts. This supports the repeatability of measurements over short monitoring periods, a prerequisite for applications such as participant selection for clinical trials. However, the utility of the score for trial enrichment remains to be established prospectively. Exploratory fairness analyses in the Verily Study Watch cohort showed high discriminative performance for the PD versus HC classification task across evaluable age and sex subgroups, with minor to no disparities. The largest disparities, both in error rates (EOD = 0.33) and in discrimination, were observed between BMI subgroups, with a drop in AUROC for the normal weight subgroup compared to the overweight subgroup (|ΔAUROC| = 0.13); a modest error-rate disparity was also observed for age subgroups (EOD = 0.20). However, due to the small number of HCs in certain subgroups, these findings necessitate further investigation in larger cohorts. Evaluation by race and ethnicity was not possible due to the limited diversity in the cohort.

Turning to pathological hallmark detection, the SAM-PD model effectively detected the presence of dopaminergic deficit and α-synuclein pathology in the Verily Study Watch at-risk group and demonstrated weak-to-moderate but significant and plausible correlations with the corresponding biological markers. The score performed comparably to the MDS prodromal PD risk score across pathological hallmark detection scenarios, despite the latter requiring specialized evaluation and, for certain markers, resource-intensive confirmatory testing, such as PSG. Overall, SAM-PD was consistently more sensitive than the MDS score across scenarios, at matched or slightly higher specificity. These findings align with those reported in the study by Schalkamp et al. (2025) [15]. However, across corresponding pathological hallmark detection scenarios, SAM-PD achieved higher recall and precision than the digital score reported in that study at the decision threshold maximizing the F1 score (Supplementary Tables 7 and 12). Although direct comparison is limited by differences in the study cohorts, cross-validation scheme, and potential variations in abnormal DAT scan classification, the passively captured SAM-PD score alone performed comparably in detecting dopaminergic deficit (AUROC = 0.86) to a recently reported model trained solely on smartphone motor test-derived features to directly predict DAT scan results (AUROC = 0.82; sex-adjusted AUROC = 0.80) [10].

Regarding its alignment with the established research criteria, the SAM-PD score correlated moderately with the comprehensive MDS prodromal PD risk score, and showed a weak, non-significant correlation with the mini-MDS risk score. Combining the SAM-PD and comprehensive MDS scores into an Ensemble PD score improved AUROC and AUPRC across pathological hallmark scenarios. At operating thresholds balancing sensitivity and specificity, the ensemble score achieved higher sensitivity relative to the comprehensive MDS score alone while maintaining a specificity of ≥ 0.80. These improvements in discrimination, together with the modest correlation between the scores, suggest that SAM-PD captures distinct aspects of nocturnal sleep and daily activity dynamics that may provide complementary information beyond that captured by current MDS criteria. Notably, combining the SAM-PD score with the mini-MDS score, which comprises only markers that do not require specialized testing, achieved discrimination comparable to the SAM-PD and the comprehensive MDS scores alone. Combining wearable-based passive sleep and activity tracking with a questionnaire assessing risk markers that can be self-reported with relative confidence (e.g., via a mobile application) may create a transferable, tiered screening strategy that could help triage individuals for specialized clinical assessment. Such a tiered approach is essential as questionnaire-based MDS criteria alone often lack sufficient predictive value for general population screening [9]. Moreover, unlike active digital assessments, passive monitoring enables continuous data collection with minimal user engagement. This is particularly relevant for long-term monitoring of individuals who may not yet experience symptoms and therefore have little motivation to engage with task-based protocols over extended periods to detect subtle changes [11].

Several limitations of this study should be acknowledged. Regarding the representativeness of the training sample, the class imbalance between PwPD and HC (approximately 84% positive) does not reflect population prevalence. Because probability calibration was performed using a stratified subset of this prevalence-enriched cohort, recalibration on population-representative samples will be required to better align probabilities with true disease likelihoods. In addition, Platt scaling was chosen over non-parametric calibration partly because of the small size of the held-out calibration set, which limits the precision of the calibration estimates. The small number of HCs also limits the precision of specificity estimates. Moreover, model fairness cannot be reliably assessed across underrepresented demographic groups, necessitating evaluation in broader, more diverse cohorts.

There are also certain methodological constraints that should be noted. For the PwPD and at-risk groups, DAT imaging was performed up to three years before wearable data collection. Although serial imaging shows that nigrostriatal dopaminergic decline during the prodromal phase progresses gradually over the years [30], the three-year interval may introduce disagreement between measured biomarker status and biological status during wearable monitoring. For HCs, baseline scans were retained regardless of acquisition date, assuming that the gradual age-related decline in striatal DAT binding seen in healthy individuals would not change their normal scan classification. Moreover, the gait detector used in the upper-limb movement feature extraction pipeline was trained on data from healthy individuals. Although it was optimized for high sensitivity in detecting gait bouts, further validation in populations with impaired gait patterns is warranted. Lastly, another limitation is the lack of information about the wrist on which participants wore the wearable device in the Verily Study Watch cohort, which may have obscured lateralized motor symptoms.

Overall, this study represents a meaningful advance toward integrating passive wearable-based monitoring into PD screening pathways. Further validation in larger, population-representative cohorts, such as the UK Biobank [31] and the All of Us Research Program [32] is now needed to address the limitations of the study and further establish whether the SAM-PD score and its associated sleep and activity metrics can predict conversion from at-risk status to clinically manifest PD. Such validation would extend the present cross-sectional findings to longitudinal clinical outcomes. If validated across diverse populations, combining passive wearable monitoring with established clinical risk frameworks could enable scalable identification of individuals who warrant specialist assessment and further confirmatory testing for PD.

## Methods

### Datasets and cohorts

We utilized data from the Parkinson’s Progression Markers Initiative (PPMI) Verily Study Watch sub-study (NCT01141023) and the AI-PROGNOSIS Digital Biomarkers Development (dBM-DEV) study (NCT06444789). Participants in the Verily Study Watch cohort belonged to one of three groups: (a) clinically diagnosed PwPD, including participants from the PPMI de novo PD and genetic PD cohorts, (b) HCs with no neurological conditions, or (c) individuals at risk of PD, belonging to the PPMI Prodromal cohort, with one or more of the following characteristics: one or more PD-associated genetic variants (in this cohort GBA and/or LRRK2), hyposmia according to the PPMI Consensus Cohort Committee, or PSG-confirmed RBD [33, 34]. The Verily Study Watch substudy was conducted in the U.S. from 2018 to 2020, and participants were monitored using the Verily Study Watch (Verily Life Sciences LLC, Dallas, TX, USA). The device is equipped with multiple sensors, including a triaxial accelerometer and gyroscope, photoplethysmography, ambient temperature, pressure, electrodermal activity, and humidity sensors. Participants in the dBM-DEV study cohort likewise belonged to one of three groups: (a) clinically diagnosed PwPD, (b) HCs with no neurological conditions or parasomnias, or (c) individuals at risk of PD based on PSG-confirmed RBD. The dBM-DEV study was conducted in Germany, France, and Spain between October 2024 and January 2026, and participants were monitored using the Garmin Vivoactive 5 smartwatch (Garmin Ltd., Schaffhausen, Switzerland). The device is equipped with a triaxial accelerometer and photoplethysmography.

In both cohorts, we applied wrist-wearable wear-time inclusion criteria requiring at least one 14-day monitoring window with a minimum of seven valid days and seven valid nights. A day was considered valid if the participant wore the device for at least 10 hours during the waking period [35], defined in this study as 06:00–23:00. A night was considered valid if the sleep detection algorithm (‘Sleep and activity metrics extraction’, Methods) identified a main sleep block of at least four hours. After applying the wear-time inclusion criteria, 283 participants from the Verily Study Watch cohort (133 PwPD, 26 HCs, 124 at-risk individuals) and 61 participants from the dBM-DEV study cohort (20 PwPD, 20 HCs, 21 at-risk individuals) qualified for further analysis. For those participants, all non-overlapping 14-day windows meeting the minimum requirement of seven valid days and seven valid nights during the observation period were included in the analysis.

The PPMI study and Verily Study Watch substudy were approved by the relevant institutional review boards or ethics committees at participating sites, and all participants provided written informed consent. The PPMI study is registered at ClinicalTrials.gov (NCT01141023). This study involved secondary analysis of de-identified data obtained from the PPMI database (https://www.ppmi-info.org) and additionally used DAT scan and CSF α-synuclein SAA results from the at-risk group, obtained also from the PPMI database after approval by the PPMI Data Access Committee. The PPMI clinico-demographic data were accessed on 12 March 2024, the genetic data on 16 April 2024, the Verily Study Watch sensor data on 22 November 2023, and the sequestered biomarker data (DAT scan and CSF α-synuclein SAA) on 19 November 2024. The AI-PROGNOSIS dBM-DEV study was approved by the relevant ethics committees at participating sites (Germany: Ethikkommission an der TU Dresden – BO-EK-2012024; France: Comité de protection des personnes Ile de France III – 2023-A02136-59; Spain: Comité de Ética de la Investigación con medicamentos del Hospital Ruber Internacional de Madrid – Sa-18773 / 15 - EC: 535), and all participants provided written informed consent. The AI-PROGNOSIS dBM-DEV study is registered at ClinicalTrials.gov (NCT06444789). This study involved secondary analysis of de-identified data obtained from the AI-PROGNOSIS dBM-DEV study database on 2 July 2026.

### Missing data handling

Height and weight were missing for five dBM-DEV participants (three at-risk, one PwPD, one HC). Values were imputed using sex- and age-specific national reference means [36] for four participants and sex-specific dBM-DEV cohort means for the remaining one. Of the five participants, one at-risk and one HC were among the 61 participants included in the analysis.

### Sleep and activity metrics extraction

For this study, we focused exclusively on accelerometer data, as it is the most ubiquitous sensor in wrist wearables. Raw tri-axial accelerometer signals were retrieved, and segments affected by sensor malfunctions, battery depletion, or other acquisition errors were excluded. Data acquired by both the Verily Study Watch, at a nominal sampling frequency of 100 Hz, and the Garmin Vivoactive 5, at a nominal sampling frequency of 25 Hz, were expressed in g and resampled to a uniform frequency of 25 Hz. Linear interpolation was used in the sleep and ambulatory metric extraction pipelines, whereas polyphase anti-aliasing resampling was used in the upper-limb movement metric extraction pipeline. Using accelerometer data sampled at 25 Hz facilitates reproducibility and consistent downstream analyses, as this is a typical sampling frequency for consumer-grade wrist wearables.

From the preprocessed signals, we estimated sleep and activity metrics. Activity metrics comprised upper-limb movement and ambulatory metrics. These metrics were selected to quantify aspects of motor function and sleep that may be affected by PD, even in its early stages. Sleep metrics were computed from the preprocessed acceleration signals using the open-source sleep detection and sleep staging pipeline ‘asleep’, developed by the Oxford Wearables group [37]. Its core human activity recognition model has been validated against video recordings and diary logs [38, 39], and its sleep staging model against polysomnography [37]. The pipeline identifies sleep windows and computes within-sleep metrics, including total sleep time, sleep stage duration, and sleep fragmentation. We considered only nighttime sleep periods and all sleep metrics were computed at a daily resolution. Ambulatory metrics, including number of steps and cadence, were derived using the open-source ‘stepcount’ pipeline, also developed by the Oxford Wearables group [40], which detects ambulatory activity and steps from wrist acceleration [40]. Its core step detection model has been validated against video recordings [40]. Metrics were computed at minute-level or hourly-level resolution.

Upper-limb movement metrics included hourly acceleration, computed as the Euclidean norm minus one (ENMO) of the triaxial acceleration signal [41], and kinematic features derived from a previously-validated pipeline for assessing movement slowness [42]. Briefly, this pipeline identifies five-second intervals of hand movement from triaxial acceleration data, excludes bouts classified as gait, and computes kinematic features from the acceleration vector magnitude within the remaining bouts, which are then statistically aggregated over 10-minute periods. For the gait detector, we trained a LightGBM classifier using time- and frequency-domain features extracted from the acceleration vector magnitude in the CAPTURE-24 dataset (n = 151 participants) [43]. The classifier achieved an AUROC of 0.78 (95% CI: 0.76–0.79), evaluated under LOSO cross-validation. We set the operating threshold to 0.20 to prioritize gait detection sensitivity, thereby minimizing the risk of undetected gait periods contaminating the kinematic features of interest. This flexibility in tuning the operating threshold, combined with the five-second granularity of hand-movement segmentation, necessitated a custom gait detector rather than the ‘stepcount’ algorithm used for extracting the ambulatory metrics.

All computed sleep and activity metrics are summarized in Supplementary Table 2. Finally, all sleep and activity metrics were temporally aggregated per participant by computing summary statistics. For each metric, we calculated the mean, median, standard deviation (SD), 25th percentile (Q25), and 75th percentile (Q75). Unless otherwise stated, temporal aggregation was performed by pooling the valid days and nights of each participant’s eligible 14-day monitoring windows over the entire observation period and computing each statistic once on the pooled data.

### Machine learning

We developed the SAM-PD model by training a classifier to differentiate between PwPD and HCs using the temporally-aggregated sleep and activity metrics from the Verily Study Watch cohort. Model development was carried out in two stages. First, we employed a nested leave-one-subject-out (LOSO) cross-validation to perform feature selection, optimize hyperparameters, calibrate predicted probabilities, and evaluate candidate classifiers. Second, we trained and calibrated the final SAM-PD model using the consensus feature set, best-performing classifier, and hyperparameter configuration derived across the LOSO folds for subsequent application to the Verily Study Watch at-risk group and the dBM-DEV cohort.

During the first stage, we applied a two-step feature selection procedure within the outer training set of each LOSO fold to retain metrics that were both temporally reliable and not highly correlated. In the first step, we assessed the test-retest reliability of each candidate metric aggregated over 14 days, using the Intraclass Correlation Coefficient (ICC) (Test-retest reliability analysis, Methods). Metrics with an ICC(2,1) 95% CI lower bound below 0.75 were excluded to minimize the influence of noisy measurements on downstream model performance. Second, to address multicollinearity, we aggregated the remaining sleep and activity metrics over the entire observation period, identified clusters of highly correlated metrics (Spearman |ρ| ≥ 0.90), and retained the most temporally reliable metric from each cluster.

Following feature selection, the participants of the outer training set were group-stratified and split into an 80% training subset for model fitting and hyperparameter optimization and a 20% calibration subset for probability calibration. Fitting and calibration were performed using the participants’ selected sleep and activity metrics, aggregated over the entire observation period. Three classification algorithms were evaluated for the SAM-PD model: ridge logistic regression, balanced random forest, and gradient-boosted trees (LightGBM). Hyperparameters were optimized within the training subset using an exhaustive grid search over predefined configurations (Supplementary Table 13). Each configuration was evaluated using stratified group-based 5-fold cross-validation, repeated five times, with AUROC as the optimization metric. Predicted probabilities were calibrated on the calibration subset using Platt scaling, which was selected because its low complexity is better suited to the small calibration-set size. Class imbalance was handled intrinsically by the balanced random forest, whereas class weighting was applied to the other classifiers. The classifiers were evaluated on the held-out participants to obtain subject-level performance estimates for the PwPD-versus-HC classification task.

In the second stage, we trained and calibrated the classifier that achieved the highest AUROC and lower ECE after calibration in the first stage, using data from all PwPD and HC participants and following the same training-calibration split strategy. We used the consensus feature set and hyperparameter configuration derived across the LOSO folds. The consensus feature set comprised metrics selected in more than half of the LOSO folds. For numerical and categorical hyperparameters, the consensus value was the median and mode across folds, respectively. The resulting SAM-PD model was then evaluated, without any retraining or recalibration, on the Verily Study Watch at-risk group and the dBM-DEV study cohort. Unless otherwise stated, SAM-PD scores for participants in these two groups correspond to the probabilities generated by the final trained and calibrated SAM-PD model from their temporally-aggregated sleep and activity metrics. In contrast, SAM-PD scores for PwPD and HCs in the Verily Study Watch cohort correspond to the test-subject probabilities obtained during the LOSO cross-validation of the best-performing calibrated classifier.

### Evaluation against pathological hallmarks

We assessed the ability of the SAM-PD, MDS prodromal PD risk, and Ensemble PD scores to detect dopaminergic deficit and α-synuclein pathology. Specifically, we first evaluated the correlation of each score with continuous biomarker measurements derived from DAT scan imaging and CSF α-synuclein SAA. We then assessed each score’s ability to distinguish individuals with normal and abnormal test results. Analyses were performed in the Verily Study Watch cohort.

### Dopaminergic deficit

For the participants in the Verily Study Watch cohort, dopaminergic deficit was assessed using DAT scan imaging before the wearable data collection period. To account for disease progression between imaging and wearable monitoring, we excluded DAT scans acquired more than three years before the wearable data collection in the PwPD and at-risk cohorts [44]. For HCs, we retained all DAT scans regardless of their acquisition date, as no meaningful progression of dopaminergic deficiency is expected in this cohort. For each participant, we quantified dopaminergic deficit using the minimum putaminal Specific Binding Ratio (SBR) from the DAT scan closest to wearable data collection. The putaminal SBR of the most affected hemisphere is the most discriminative single DAT-derived measure, as nigrostriatal degeneration preferentially begins in the posterior putamen during the premotor stage [45]. For the classification analysis, participants were categorized as having normal dopaminergic function or dopaminergic deficit based on their DAT scan result. DAT scan results were classified as normal or abnormal according to the PPMI visual interpretation and consensus process. Each scan was independently rated by two reviewers, with a third reviewer adjudicating discordant ratings; the consensus classification was determined by agreement between two reviewers. When no visual interpretation was available, scans were classified as abnormal if the minimum putaminal SBR was below 1.037, the PPMI-derived threshold for distinguishing PwPD from HCs, and as normal otherwise [46]. Scans were classified by visual interpretation in all 26 HCs, 28 of 86 PwPD, and 56 of 121 at-risk participants, whereas the remaining scans were classified by the SBR threshold. Because DAT scan imaging preceded wearable data collection, the dopaminergic-deficit analysis is cross-sectional with respect to the SAM-PD and MDS scores and should be interpreted as assessing the association between the score and pre-existing dopaminergic status rather than the prediction of a future outcome.

### α-Synuclein pathology

α-synuclein SAA data were available for participants who underwent lumbar puncture for cerebrospinal fluid collection [47]. CSF was collected before wearable data collection, and SAA assays were performed on the stored samples afterward. The assay was performed using two protocols, with fluorescence read at 24 or 150 hours. Because maximum fluorescence is protocol-dependent, correlation analyses were restricted to the 150-hour readout, which was the only protocol applied across the entire at-risk cohort. For these analyses, we used the mean maximum fluorescence (Fmax) of three replicates. For the classification analysis, participants were categorized as having α-synuclein pathology based on a positive SAA result or not having α-synuclein pathology based on a negative SAA result, regardless of assay protocol. We used the SAA result from the CSF sample collected closest to, and before, the start of wearable data collection. If the sample was assayed more than once, the earliest assay result was used.

### Presence of either or both pathological hallmarks

We further evaluated the ability of SAM-PD and MDS scores to identify individuals exhibiting at least one or both pathological hallmarks. For the at-least-one-hallmark analysis, participants with an abnormal result on either test were considered positive, even if the result of the other test was unavailable. In the both-hallmark analysis, participants were considered positive if they had an abnormal DAT scan and a positive SAA result. Participants with neither pathological feature, defined as having both DAT scan and SAA results available and negative, served as the comparison group in both analyses.

### MDS prodromal PD risk score estimation and combination strategies

We first compared the SAM-PD score with the MDS prodromal PD risk score and then investigated their complementary ability to detect dopaminergic deficit and α-synuclein pathology in the Verily Study Watch cohort. The MDS prodromal PD risk score is the output of a Bayesian classifier that estimates the probability of prodromal PD based on established risk markers and prodromal features [7]. It first assigns an age-based pretest probability, which is then updated using Bayesian likelihood ratios for each risk marker or prodromal feature present. Likelihood ratios have been derived from epidemiological studies [8].

We computed two versions of the MDS prodromal PD risk score: a version utilizing all available risk and prodromal marker information and a version relying only on markers that do not require specialized ancillary testing and can be self-reported with relative confidence, referred to as the mini-MDS risk score. The former operationalized the risk and prodromal markers of the MDS research criteria for prodromal PD that were available in the PPMI dataset, comprising 23 variables. These included, but were not limited to, the polygenic risk score (PRS) or presence of genetic mutations, such as those in the GBA or LRRK2 genes, olfactory loss confirmed by the University of Pennsylvania Smell Identification Test (UPSIT), subthreshold parkinsonism assessed using MDS-UPDRS Part III, and PSG-confirmed RBD. The omitted markers were the DAT scan result, which served as one of the ground truths of this study; substantia nigra hyperechogenicity, occupational solvent exposure and plasma urate, as these data were not uniformly collected in PPMI; and physical inactivity, for which no relevant assessment or measure was available. In addition to age, the mini-MDS risk score was calculated using the following variables, adopting an approach similar to De Klerk et al. [9]: biological sex, smoking status (never, current, former), caffeine consumption, having a first-degree relative with PD, RBD screening questionnaire result, and the presence of type II diabetes mellitus, constipation, daytime somnolence, urinary dysfunction, depression, and cognitive impairment. A complete list of all markers used in both versions of the MDS risk score is provided in Supplementary Table 14. All markers were extracted from assessments conducted before or at the beginning of the Verily Study Watch data collection. Former smoking status, caffeine consumption, and occupational exposure to pesticides reflect historical exposure.

To investigate their complementary ability to detect pathological hallmarks, we evaluated three approaches for combining the SAM-PD and MDS scores: maximum score-based screening, sequential screening, and score fusion using a meta-learner. In the maximum score approach, participants were classified as positive (having one, either, or both pathologies) if either score exceeded its respective decision threshold. In the sequential screening approach, participants were classified as positive only if both scores exceeded their respective decision thresholds. For the meta-learner approach, we trained a logistic regression model to distinguish PwPD from HCs using the SAM-PD and MDS scores as predictors. Logistic regression was selected as the meta-learner because of its simplicity and interpretable weighting of the input predictions. The meta-learner was trained on the out-of-fold SAM-PD probabilities and MDS scores of the Verily Study Watch PwPD and HC participants, and its performance and operating thresholds were estimated with nested LOSO cross-validation. In each outer fold, the SAM-PD probabilities used to train the meta-learner were regenerated with the held-out participant excluded from SAM-PD training, ensuring that the participant’s data were not used to train either model before the final prediction was generated. Similar to the SAM-PD model, hyperparameters for the logistic regression model were optimized using an exhaustive grid search over predefined configurations within this scheme. Each configuration was evaluated using stratified group 5-fold cross-validation, repeated three times, with AUROC as the optimization metric. The final logistic regression model was then trained on the complete PwPD and HC dataset using the hyperparameter configuration derived across the LOSO folds. This final logistic regression meta-learner, together with the MDS risk score and the SAM-PD model trained and calibrated on the entire PwPD and HC dataset, formed the final system. This system was applied without further tuning to the Verily Study Watch at-risk group to generate Ensemble PD scores. All combination strategies were evaluated using both the MDS and mini-MDS risk scores; for the meta-learner approach, the latter yielded the mini-Ensemble PD score.

### Polygenic risk score calculation

PRSs were calculated for each participant in the Verily Study Watch cohort. The PRS calculation was based on the combined set of single nucleotide polymorphisms (SNPs) associated with PD identified in the meta-analyses by Nalls et al. [48] and Leonard et al. [49]. Ambiguous, multiallelic, and variants located in the LRRK2 gene were systematically excluded from the analysis. Each PRS was calculated as a weighted sum of the previously identified risk alleles, following the methodology described by Mavaddat et al. [50], using the formula:

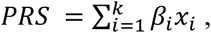

where *β_i_* = ln(*OR_i_*), *OR_i_* is the odds ratio (OR) of each SNP *i* reported in the corresponding GWAS meta-analysis, *x_i_* ∈ {0,1,2} is the dosage of the risk allele for SNP *i*, and *k* is the total number of SNPs included in the PRS.

For inclusion in the MDS prodromal PD risk score, participants were subsequently classified into high-or low-PRS categories using quartile thresholds derived from the PRS distribution in the unified Accelerating Medicines Partnership Parkinson’s disease (AMP-PD) cohorts [51]. Participants with PRS values within the interquartile range were considered unclassified, and the PRS-associated marker was not included in the calculation of the MDS score.

### Classification performance evaluation

Classification performance was evaluated using AUROC, AUPRC, sensitivity, specificity, precision, balanced accuracy, and F1 score. Because the classification tasks investigated in this study exhibited varying degrees of class imbalance, we used both AUROC and AUPRC as primary evaluation metrics. For the SAM-PD and Ensemble PD scores, operating point-dependent metrics were estimated at the Youden-optimal threshold, derived from the corresponding PwPD-versus-HC classification tasks in the Verily Study Watch cohort. During LOSO cross-validation, fold-specific thresholds were used, the median of these thresholds was applied to the at-risk group and the dBM-DEV cohort. The Verily Study Watch at-risk group and dBM-DEV cohort were not used for threshold selection. Performance was also evaluated at two alternative operating points representing plausible screening configurations: the threshold that maximized the F1 score and the threshold that maximized sensitivity while maintaining specificity ≥ 0.50. For all versions of the MDS risk score, the recommended threshold for prodromal PD (0.80) was used [8].

### Fairness analysis

To assess potential demographic disparities in SAM-PD model performance, we stratified the PwPD and HC predictions of the Verily Study Watch cohort by age, sex, BMI, ethnicity, and race. Within each subgroup, classification performance was evaluated using AUROC and AUPRC, calculated only for subgroups with at least five participants in each outcome class. Race and ethnicity subgroups were defined according to PPMI. For age subgrouping, we followed the ranges defining young-onset (< 50 years), middle-onset (50-69 years), and late-onset (≥70 years) PD [52]. For BMI subgrouping, we followed the World Health Organization categories [53]: underweight (BMI < 18.5), normal weight (25.0 > BMI ≥ 18.5), overweight (30.0 > BMI ≥ 25.0), and obesity (BMI ≥ 30.0). We also assessed disparities using three metrics: the Demographic Parity Difference (DPD), Demographic Parity Ratio (DPR), and Equalized Odds Difference (EOD).

### Test-retest reliability analysis

We performed a test-retest reliability analysis to determine the minimum temporal aggregation interval to obtain reliable SAM-PD scores and constituent sleep and activity metrics. We followed an approach similar to [54]. The analysis covered each participant’s first qualifying fortnight and the immediately subsequent fortnight, assuming each participant’s health status remained fairly stable throughout this interval. Sleep and activity metrics were aggregated over non-overlapping windows of 3, 7, 10, and 14 days and used to derive the corresponding SAM-PD scores. A calendar day was considered valid if the participant wore the wearable device for at least 10 hours during the 06:00–23:00 waking period and a main sleep block of at least four hours was detected for the corresponding night. For each aggregation interval, the aggregated metrics and resulting SAM-PD scores from the first and second windows were treated as test and retest measurements, respectively. For each aggregation interval, we calculated the two-way random effects, absolute agreement, single-measurement intraclass correlation coefficient [ICC(2,1)] with 95% confidence intervals [20] for all metrics and the SAM-PD score. Only participants with at least one valid test-retest pair at a given aggregation interval were included in the corresponding ICC analysis.

### Model interpretability analysis

To characterize how each input contributed to the SAM-PD score, we computed SHAP values [55] on held-out data within the LOSO cross-validation. In each LOSO fold, SHAP values for the held-out participant were computed with the same fitted model that generated their prediction. SHAP values were estimated with the permutation explainer applied to the calibrated SAM-PD probability. Individual feature importance was quantified as the mean absolute SHAP value across participants.

### Statistical analyses

Demographic characteristics were compared between groups within each cohort and between cohorts within each group using two-sided Mann-Whitney U tests for continuous variables (age and BMI) and Pearson’s chi-squared tests without continuity correction for sex. No multiple-testing correction was applied to these baseline comparisons because they were intended as descriptive comparisons. Pairwise comparisons of the age- and sex-accounting SAM-PD, MDS, and Ensemble PD scores were performed using two-sided Mann-Whitney U tests, whereas comparisons of individual sleep and activity metrics were performed using rank-based analysis of covariance (ANCOVA) adjusted for age and sex [56]. Statistical significance was assessed at a nominal threshold of α = 0.05. Bonferroni correction was applied for multiple comparisons, where indicated. Unless otherwise stated, continuous variables are reported as mean ± standard deviation, and categorical variables as n (%). Pearson’s correlation coefficients were calculated to assess linear relationships between continuous variables. Spearman rank correlation was used for feature selection to account for potential non-linear associations. For ICC analyses, CIs were derived using the F-distribution. To quantify uncertainty in classification performance, 95% CIs for sensitivity, specificity and precision were estimated using the Clopper-Pearson method [57], whereas 95% CIs for all other performance metrics were calculated using 1,000 bootstrap iterations with replacement. Analyses were conducted in Python 3.10.9. Raw accelerometry was processed with actipy 3.8.0, sleep and step metrics were derived with asleep 0.4.16 and stepcount 3.9.1 (self-supervised model, weights ssl-20230208), respectively, and temporal aggregation statistics were computed with tsfresh 0.20.2. Machine learning models were implemented using scikit-learn 1.1.1, imbalanced-learn 0.9.1, and LightGBM 3.3.0, and SHAP analysis was performed using shap 0.42.1. A fixed random seed of 42 was used for all stochastic procedures. Statistical testing was performed using scipy 1.10.1, pingouin 0.5.4, and statsmodels 0.14.2. Visualizations were generated using seaborn 0.12.2 and matplotlib 3.7.5. PRS calculations were performed in R 4.3.2. All analyses, except PRS calculations, were performed on a workstation equipped with a 24-core CPU (3.8 GHz), 128 GB of RAM, and 48 GB of VRAM.

## Code availability

Code that supports the findings of this study will be made available on GitHub at https://github.com/spbtu/sam-pd-paper upon publication.

## Data Availability

Parkinson’s Progression Markers Initiative (PPMI) data used in this study were obtained from the study database (www.ppmi-info.org/access-data-specimens/download-data, RRID: SCR 006431). Access to PPMI data requires registration and formal request approval. At the time of writing, access to dBM-DEV data may be granted upon reasonable request to the corresponding authors and approval by the dBM-DEV study data controllers.

## Acknowledgements

This work was supported by the AI-PROGNOSIS project funded by the European Union under Grant Agreement No. 101080581. Views and opinions expressed are those of the authors only and do not necessarily reflect those of the European Union or European Health and Digital Executive Agency. Neither the European Union nor the European Health and Digital Executive Agency can be held responsible for them. Data used in the preparation of this article were obtained from the Parkinson’s Progression Markers Initiative (PPMI) database. For up-to-date information on the study, visit www.ppmi-info.org. PPMI—a public-private partnership—is funded by the Michael J. Fox Foundation for Parkinson’s Research and funding partners, including 4D Pharma, Abbvie, AcureX, Allergan, Amathus Therapeutics, Aligning Science Across Parkinson’s, AskBio, Avid Radiopharmaceuticals, BIAL, BioArctic, Biogen, Biohaven, BioLegend, BlueRock Therapeutics, Bristol-Myers Squibb, Calico Labs, Capsida Biotherapeutics, Celgene, Cerevel Therapeutics, Coave Therapeutics, DaCapo Brainscience, Denali, Edmond J. Safra Foundation, Eli Lilly, Gain Therapeutics, GE HealthCare, Genentech, GSK, Golub Capital, Handl Therapeutics, Insitro, Jazz Pharmaceuticals, Johnson & Johnson Innovative Medicine, Lundbeck, Merck, Meso Scale Discovery, Mission Therapeutics, Neurocrine Biosciences, Neuron23, Neuropore, Pfizer, Piramal, Prevail Therapeutics, Roche, Sanofi, Servier, Sun Pharma Advanced Research Company, Takeda, Teva, UCB, Vanqua Bio, Verily, Voyager Therapeutics, the Weston Family Foundation, and Yumanity Therapeutics.

## Author contributions

A.M., I.G., C.S. and S.H. designed the study. A.M. performed the machine learning and statistical analysis with the support of I.G. I.G. performed the MDS prodromal PD risk score calculations. S.H. provided expertise input on the analysis. P.C. performed the PRS calculations with the support of K.M. and K.C. N.D.C., T.F., N.S., M.-L.A.M., M.K., M.F., and B.F. contributed to the dBM-DEV study data collection.

A.M. wrote the initial draft of the manuscript. S.H., C.S., and I.G. edited the manuscript. M.K., T. F., M.F., and B.F. provided clinical expertise and contributed to the interpretation of the results. Figures and tables were done by A.M. with the support of S.H. L.H. critically reviewed the manuscript. All authors read and approved the manuscript.

## Corresponding authors

Correspondence and requests for materials should be addressed to Apostolos Moustaklis, Leontios Hadjileontiadis or Stelios Hadjidimitriou.

