## Supplementary Table 1 for "Wearable-based sleep and activity tracking detects Parkinson’s disease and associated pathological hallmarks"

### **Supplementary Tables**

| **Comparison** | **Variable** | **Group 1** | **Group 2** | **Test** | **p value** |
| --- | --- | --- | --- | --- | --- |
| Verily Study Watch: PwPD (n = 133) versus HC (n = 26) | Females, n (%) | 54 (40.6%) | 12 (46.2%) | Pearson’s chi-squared | 0.599 |
| Verily Study Watch: PwPD (n = 133) versus HC (n = 26) | Age (years) | 69.0 ± 7.9 | 69.1 ± 11.1 | Mann-Whitney U | 0.782 |
| Verily Study Watch: PwPD (n = 133) versus HC (n = 26) | BMI (kg/m²) | 26.9 ± 4.9 | 26.7 ± 4.6 | Mann-Whitney U | 0.736 |
| Verily Study Watch: At-risk (n = 124) versus HC (n = 26) | Females, n (%) | 76 (61.3%) | 12 (46.2%) | Pearson’s chi-squared | 0.154 |
| Verily Study Watch: At-risk (n = 124) versus HC (n = 26) | Age (years) | 65.9 ± 7.1 | 69.1 ± 11.1 | Mann-Whitney U | 0.026 |
| Verily Study Watch: At-risk (n = 124) versus HC (n = 26) | BMI (kg/m²) | 27.7 ± 5.4 | 26.7 ± 4.6 | Mann-Whitney U | 0.370 |
| Verily Study Watch: PwPD (n = 133) versus At-risk (n = 124) | Females, n (%) | 54 (40.6%) | 76 (61.3%) | Pearson’s chi-squared | < 0.001 |
| Verily Study Watch: PwPD (n = 133) versus At-risk (n = 124) | Age (years) | 69.0 ± 7.9 | 65.9 ± 7.1 | Mann-Whitney U | < 0.001 |
| Verily Study Watch: PwPD (n = 133) versus At-risk (n = 124) | BMI (kg/m²) | 26.9 ± 4.9 | 27.7 ± 5.4 | Mann-Whitney U | 0.297 |
| dBM-DEV: PwPD (n = 20) versus HC (n = 20) | Females, n (%) | 10 (50.0%) | 6 (30.0%) | Pearson’s chi-squared | 0.197 |
| dBM-DEV: PwPD (n = 20) versus HC (n = 20) | Age (years) | 62.1 ± 6.4 | 64.3 ± 10.3 | Mann-Whitney U | 0.481 |
| dBM-DEV: PwPD (n = 20) versus HC (n = 20) | BMI (kg/m²) | 27.7 ± 8.2 | 25.7 ± 3.5 | Mann-Whitney U | 0.735 |
| dBM-DEV: At-risk (n = 21) versus HC (n = 20) | Females, n (%) | 5 (23.8%) | 6 (30.0%) | Pearson’s chi-squared | 0.655 |
| dBM-DEV: At-risk (n = 21) versus HC (n = 20) | Age (years) | 71.5 ± 5.9 | 64.3 ± 10.3 | Mann-Whitney U | 0.049 |
| dBM-DEV: At-risk (n = 21) versus HC (n = 20) | BMI (kg/m²) | 26.0 ± 2.9 | 25.7 ± 3.5 | Mann-Whitney U | 0.784 |
| dBM-DEV: PwPD (n = 20) versus At-risk (n = 21) | Females, n (%) | 10 (50.0%) | 5 (23.8%) | Pearson’s chi-squared | 0.082 |
| dBM-DEV: PwPD (n = 20) versus At-risk (n = 21) | Age (years) | 62.1 ± 6.4 | 71.5 ± 5.9 | Mann-Whitney U | < 0.001 |
| dBM-DEV: PwPD (n = 20) versus At-risk (n = 21) | BMI (kg/m²) | 27.7 ± 8.2 | 26.0 ± 2.9 | Mann-Whitney U | 0.969 |
| HC: Verily Study Watch (n = 26) versus dBM-DEV (n = 20) | Females, n (%) | 12 (46.2%) | 6 (30.0%) | Pearson’s chi-squared | 0.266 |
| HC: Verily Study Watch (n = 26) versus dBM-DEV (n = 20) | Age (years) | 69.1 ± 11.1 | 64.3 ± 10.3 | Mann-Whitney U | 0.118 |
| HC: Verily Study Watch (n = 26) versus dBM-DEV (n = 20) | BMI (kg/m²) | 26.7 ± 4.6 | 25.7 ± 3.5 | Mann-Whitney U | 0.618 |
| At-risk: Verily Study Watch (n = 124) versus dBM-DEV (n = 21) | Females, n (%) | 76 (61.3%) | 5 (23.8%) | Pearson’s chi-squared | 0.001 |
| At-risk: Verily Study Watch (n = 124) versus dBM-DEV (n = 21) | Age (years) | 65.9 ± 7.1 | 71.5 ± 5.9 | Mann-Whitney U | < 0.001 |
| At-risk: Verily Study Watch (n = 124) versus dBM-DEV (n = 21) | BMI (kg/m²) | 27.7 ± 5.4 | 26.0 ± 2.9 | Mann-Whitney U | 0.201 |
| PwPD: Verily Study Watch (n = 133) versus dBM-DEV (n = 20) | Females, n (%) | 54 (40.6%) | 10 (50.0%) | Pearson’s chi-squared | 0.427 |
| PwPD: Verily Study Watch (n = 133) versus dBM-DEV (n = 20) | Age (years) | 69.0 ± 7.9 | 62.1 ± 6.4 | Mann-Whitney U | < 0.001 |
| PwPD: Verily Study Watch (n = 133) versus dBM-DEV (n = 20) | BMI (kg/m²) | 26.9 ± 4.9 | 27.7 ± 8.2 | Mann-Whitney U | 0.543 |

**Supplementary Table 1** | Comparison of sex, age, and body mass index (BMI) between groups within each cohort and between cohorts within each group. Values are presented as number of females (percentage) or mean ± standard deviation. Age and BMI were compared using two-sided Mann-Whitney U tests and sex using Pearson's chi-squared tests. No correction for multiple testing was applied.

| **Domain** | **Metric** | **Resolution** | **Short description** | **Algorithm** |
| --- | --- | --- | --- | --- |
| Sleep | Total bed time (h) | Per sleep period (night) | Time between sleep onset and final wake, i.e. the sleep opportunity window | ‘asleep’  (Yuan et al., 2024) |
| Sleep | Sleep onset latency (min) | Per sleep period (night) | Time from going to bed to the first epoch scored as sleep |  |
| Sleep | Total sleep time (min) |  | Total time scored in any sleep stage during the sleep period |  |
| Sleep | Wake after sleep onset (min) |  | Time scored as wake between sleep onset and final wake; indicative of sleep fragmentation |  |
| Sleep | Wake duration (min) |  | Total wake time within the sleep period, including wake before sleep onset |  |
| Sleep | Sleep efficiency (%) |  | Total sleep time as a percentage of total bed time |  |
| Sleep | REM sleep latency (min) |  | Time from sleep onset to the first REM sleep bout |  |
| Sleep | REM sleep duration (min) |  | Total time scored as REM sleep |  |
| Sleep | NREM sleep duration (min) |  | Total time scored as non-REM sleep (N1 + N2 + N3), equal to total sleep time minus REM duration |  |
| Upper-limb movement | Peak acceleration magnitude median (g) | Per 10-min interval including hand-movement and no concurrent gait | Median peak acceleration magnitude across 5-s bouts of hand movement within a 10-min interval; indicative of movement vigor | Gerasimou et al., 2025 |
| Upper-limb movement | Peak acceleration magnitude 75^th^ percentile (Q75) (g) |  | Q75 of the peak acceleration magnitude across 5-s hand-movement bouts within a 10-min interval |  |
| Upper-limb movement | Peak acceleration magnitude standard deviation (SD) (g) |  | Variability of the peak acceleration magnitude across 5-s hand-movement bouts within a 10-min interval |  |
| Upper-limb movement | Short-term acceleration entropy median (bits) |  | Median of the Shannon entropy of the acceleration-magnitude distribution across 5-s hand-movement bouts within a 10-min interval; lower entropy indicates smoother short-term movement |  |
| Upper-limb movement | Short-term acceleration entropy Q75 (bits) |  | Q75 of short-term acceleration entropy across 5-s hand-movement bouts within a 10-min interval |  |
| Upper-limb movement | Short-term acceleration entropy SD (bits) |  | Variability of the short-term acceleration entropy across 5-s hand-movement bouts within a 10-min interval |  |
| Upper-limb movement | Smoothness magnitude median (g/s³) |  | Median jerkiness of movement (squared rate of change of acceleration, normalized to amplitude and duration) across 5-s hand-movement bouts within a 10-min interval; lower values reflect slower, less dynamic movement |  |
| Upper-limb movement | Smoothness magnitude Q75 (g/s³) |  | 75th percentile of movement jerkiness across 5-s hand-movement bouts within a 10-min interval |  |
| Upper-limb movement | Smoothness magnitude SD (g/s³) |  | Variability of movement jerkiness across 5-s hand-movement bouts within a 10-min interval |  |
| Upper-limb movement | Range of motion magnitude median (g) |  | Median acceleration range across 5-s hand movement bouts within a 10-min interval; a proxy for movement amplitude |  |
| Upper-limb movement | Range of motion magnitude Q75 (g) |  | 75th percentile of the acceleration range across 5-s hand-movement bouts within a 10-min interval |  |
| Upper-limb movement | Range of motion magnitude SD (g) |  | Variability of the acceleration range across 5-s hand-movement bouts within a 10-min interval |  |
| Upper-limb movement | Acceleration root mean squared (RMS) magnitude median (g) |  | Median RMS acceleration magnitude across 5-s hand-movement bouts within a 10-min interval; an overall measure of movement intensity |  |
| Upper-limb movement | Acceleration RMS magnitude Q75 (g) |  | 75th percentile of the RMS acceleration magnitude across 5-s hand-movement bouts within a 10-min interval |  |
| Upper-limb movement | Acceleration RMS magnitude SD (g) |  | Variability of the RMS acceleration magnitude across 5-s hand-movement bouts within a 10-min interval |  |
| Ambulatory activity | Hourly steps (count) | Hourly | Step count per hour; indicative of the volume of walking activity | ‘stepcount’ (Small et al., 2024) |
| Upper-limb movement | Hourly acceleration Euclidean norm minus one (ENMO) (mg) |  | Mean acceleration ENMO per hour; indicative of overall movement intensity, regardless of walking |  |
| Ambulatory activity | Cadence mean (steps/min) | Per walking bout | Mean stepping rate during detected walking bouts |  |
| Ambulatory activity | Cadence SD (steps/min) |  | Within-bout variability of stepping rate |  |
| Ambulatory activity | Cadence 25^th^ percentile (Q25) (steps/min) |  | 25th percentile of within-bout stepping rate, characterizing the slowest walking pace |  |
| Ambulatory activity | Cadence median (steps/min) |  | Median within-bout stepping rate |  |
| Ambulatory activity | Cadence 75^th^ percentile (Q75) (steps/min) |  | 75th percentile of within-bout stepping rate, characterizing the fastest walking pace |  |
| Ambulatory activity | Walking-bout acceleration ENMO (mg) |  | Mean acceleration ENMO within bout; indicative of movement intensity while walking |  |

**Supplementary Table 2 |** Sleep and activity metrics extracted from triaxial acceleration signals. The resolution column indicates the temporal resolution of each metric before aggregation over each participant’s observation period. The algorithm column specifies the algorithm used to derive each metric. Hourly acceleration (ENMO) is grouped with the upper-limb movement metrics but was computed with the ‘stepcount’ pipeline.

| **Classifier** | **AUROC (uncalibrated model)** | **AUROC (calibrated model)** | **ECE (uncalibrated model)** | **ECE  (calibrated model)** |
| --- | --- | --- | --- | --- |
| Balanced random forest (selected) | 0.92 (0.86–0.97) | 0.93 (0.88–0.97) | 0.17 (0.14–0.22) | 0.04 (0.03–0.09) |
| Ridge logistic regression | 0.90 (0.81–0.98) | 0.93 (0.88–0.97) | 0.12 (0.10–0.18) | 0.06 (0.04–0.10) |
| Gradient-boosted trees (LightGBM) | 0.89 (0.80–0.95) | 0.91 (0.85–0.97) | 0.08 (0.05–0.13) | 0.05 (0.04–0.11) |

**Supplementary Table 3 |** Performance of the classification algorithms evaluated for distinguishing PwPD (n = 133) and HCs (n = 26) in the Verily Study Watch cohort under nested LOSO cross-validation. AUROC and expected calibration error (ECE) are reported as point estimates with 95% CIs (in parentheses) from 1,000 bootstrap resamples with replacement. Performance is reported for both uncalibrated and calibrated models. Calibration was performed using Platt scaling.

| **Metric** | **Uncalibrated SAM-PD model** | **Calibrated SAM-PD model** |
| --- | --- | --- |
| AUROC (95% CI) | 0.92 (0.86–0.97) | 0.93 (0.88–0.97) |
| AUPRC (95% CI) | 0.98 (0.97–0.99) | 0.98 (0.97–1.00) |
| Brier score (95% CI) | 0.11 (0.09–0.14) | 0.07 (0.05–0.10) |
| ECE (95% CI) | 0.17 (0.14–0.22) | 0.04 (0.03–0.09) |
| Youden-optimal operating point (decision thresholds: 0.46–0.49 uncalibrated; 0.78–0.79 calibrated) | | |
| Sensitivity (95% CI) | 0.86 (0.79–0.92) | 0.89 (0.83–0.94) |
| Specificity (95% CI) | 0.81 (0.61–0.93) | 0.85 (0.65–0.96) |
| Precision (95% CI) | 0.96 (0.91–0.99) | 0.97 (0.92–0.99) |
| Balanced accuracy (95% CI) | 0.84 (0.75–0.91) | 0.87 (0.79–0.94) |
| F1 score (95% CI) | 0.91 (0.87–0.94) | 0.93 (0.89–0.96) |
| Maximum-F1 score operating point (decision thresholds: 0.17–0.28 uncalibrated; 0.41–0.60 calibrated) | | |
| Sensitivity (95% CI) | 0.97 (0.92–0.99) | 0.97 (0.92–0.99) |
| Specificity (95% CI) | 0.42 (0.23–0.63) | 0.46 (0.27–0.67) |
| Precision (95% CI) | 0.90 (0.83–0.94) | 0.90 (0.84–0.95) |
| Balanced accuracy (95% CI) | 0.70 (0.60–0.79) | 0.72 (0.62–0.81) |
| F1 score (95% CI) | 0.93 (0.90–0.96) | 0.93 (0.91–0.96) |
| Operating point maximizing sensitivity with specificity ≥ 0.50 (decision threshold: 0.28 uncalibrated; 0.47–0.60 calibrated) | | |
| Sensitivity (95% CI) | 0.97 (0.92–0.99) | 0.98 (0.94–1.00) |
| Specificity (95% CI) | 0.54 (0.33–0.73) | 0.50 (0.30–0.70) |
| Precision (95% CI) | 0.91 (0.86–0.96) | 0.91 (0.85–0.95) |
| Balanced accuracy (95% CI) | 0.75 (0.66–0.84) | 0.74 (0.63–0.83) |
| F1 score (95% CI) | 0.94 (0.91–0.97) | 0.94 (0.91–0.97) |

**Supplementary Table 4 |** Detailed performance metrics of the uncalibrated and calibrated SAM-PD models (balanced random forest) for classifying PwPD (n = 133) and HCs (n = 26) in the Verily Study Watch cohort under nested LOSO cross-validation. All metrics are reported as point estimates with 95% confidence intervals (CIs); CIs for sensitivity, specificity, and precision are exact Clopper-Pearson binomial intervals and those for all other metrics were obtained from 1,000 bootstrap resamples with replacement. Sensitivity, specificity, precision, balanced accuracy, and F1 score are reported at three operating points: the Youden-optimal point, the maximum-F1 score point, and the point that maximizes sensitivity while maintaining specificity ≥ 0.50. Each participant was classified using the decision threshold derived from the out-of-fold predictions of the remaining participants (per-fold leave-one-out approach); the ranges of the decision thresholds are shown for each operating point.

| **Feature** | **Group** | **PwPD (n)** | **HC (n)** | **AUROC** | **AUPRC** |
| --- | --- | --- | --- | --- | --- |
| Age | <50 years | 1 | 2 | — | — |
| Age | 50–69 years | 69 | 13 | 0.96 (0.91–1.00) | 0.99 (0.98–1.00) |
| Age | ≥70 years | 63 | 11 | 0.92 (0.83–0.98) | 0.98 (0.96–1.00) |
| BMI | Underweight | 3 | 1 | — | — |
| BMI | Normal weight | 48 | 9 | 0.85 (0.71–0.95) | 0.97 (0.93–0.99) |
| BMI | Overweight | 57 | 12 | 0.98 (0.94–1.00) | 1.00 (0.99–1.00) |
| BMI | Obese | 25 | 4 | — | — |
| Sex | Female | 54 | 12 | 0.96 (0.91–1.00) | 0.99 (0.98–1.00) |
| Sex | Male | 79 | 14 | 0.90 (0.80–0.97) | 0.98 (0.96–1.00) |
| Ethnicity | Ashkenazi Jewish | 44 | 2 | — | — |
| Ethnicity | Hispanic/Latino | 3 | 0 | — | — |
| Ethnicity | Not Specified | 86 | 24 | 0.94 (0.88–0.98) | 0.98 (0.96–0.99) |
| Race | American Indian/Alaska Native | 1 | 0 | — | — |
| Race | Asian | 3 | 0 | — | — |
| Race | Black | 2 | 0 | — | — |
| Race | White | 127 | 26 | 0.93 (0.87–0.97) | 0.98 (0.97–0.99) |

**Supplementary Table 5 |** Subgroup performance of the calibrated SAM-PD model for classifying PwPD and HCs in the Verily Study Watch cohort. Performance metrics are reported by age, BMI, sex, ethnicity, and race. AUROC and AUPRC are reported as point estimates with 95% confidence intervals (CIs) from 1,000 bootstrap resamples with replacement. Metrics are reported only for subgroups with at least five participants in each outcome class.

| **Protected variable** | **Demographic parity difference** | **Demographic parity ratio** | **Equalized odds difference** |
| --- | --- | --- | --- |
| Age | 0.12 (0.01–0.23) | 0.86 (0.74–0.98) | 0.20 (0.06–0.50) |
| Sex | 0.03 (0.00–0.17) | 0.97 (0.80–1.00) | 0.13 (0.02–0.42) |
| BMI | 0.00 (0.00–0.17) | 1.00 (0.80–1.00) | 0.33 (0.08–0.70) |

**Supplementary Table 6 |** Disparity metrics for the SAM-PD model across age, sex, and BMI. Demographic Parity Difference (DPD), Demographic Parity Ratio (DPR), and Equalized Odds Difference (EOD) are reported with 95% confidence intervals from 1,000 bootstrap resamples with replacement. Metrics were calculated only when at least two subgroups each had at least five participants in each outcome class; therefore, disparities were not evaluated for race and ethnicity.

| **Score** | **AUROC** | **AUPRC** | **Sensitivity** | **Specificity** | **Precision** | **Balanced accuracy** | **F1 score** |
| --- | --- | --- | --- | --- | --- | --- | --- |
| **Dopaminergic deficit** | | | | | | | |
| SAM-PD | 0.86 (0.75–0.94) | 0.45 (0.25–0.73) | 0.67 (0.38–0.88) | 0.86 (0.78–0.92) | 0.40 (0.21–0.61) | 0.76 (0.63–0.89) | 0.50 (0.27–0.67) |
| MDS | 0.80 (0.66–0.91) | 0.46 (0.21–0.68) | 0.47 (0.21–0.73) | 0.86 (0.78–0.92) | 0.32 (0.14–0.55) | 0.66 (0.52–0.80) | 0.38 (0.16–0.56) |
| mini-MDS | 0.72 (0.59–0.84) | 0.25 (0.14–0.47) | 0.07 (0.00–0.32) | 0.98 (0.93–1.00) | 0.33 (0.01–0.91) | 0.52 (0.48–0.61) | 0.11 (0.00–0.33) |
| Ensemble PD | 0.89 (0.80–0.96) | 0.61 (0.35–0.83) | 0.60 (0.32–0.84) | 0.89 (0.81–0.94) | 0.43 (0.22–0.66) | 0.74 (0.59–0.88) | 0.50 (0.26–0.67) |
| mini-Ensemble PD | 0.86 (0.76–0.94) | 0.46 (0.23–0.71) | 0.80 (0.52–0.96) | 0.81 (0.72–0.88) | 0.38 (0.21–0.56) | 0.81 (0.68–0.91) | 0.51 (0.29–0.67) |
| Maximum-score screening | — | — | 0.73 (0.45–0.92) | 0.75 (0.66–0.83) | 0.30 (0.16–0.47) | 0.74 (0.60–0.86) | 0.42 (0.23–0.58) |
| Sequential screening | — | — | 0.40 (0.16–0.68) | 0.96 (0.91–0.99) | 0.60 (0.26–0.88) | 0.68 (0.55–0.82) | 0.48 (0.20–0.69) |
| **α-Synuclein pathology** | | | | | | | |
| SAM-PD | 0.76 (0.62–0.90) | 0.50 (0.30–0.75) | 0.63 (0.38–0.84) | 0.87 (0.79–0.93) | 0.50 (0.29–0.71) | 0.75 (0.64–0.87) | 0.56 (0.36–0.72) |
| MDS | 0.76 (0.65–0.87) | 0.42 (0.25–0.63) | 0.42 (0.20–0.67) | 0.86 (0.78–0.93) | 0.38 (0.18–0.62) | 0.64 (0.53–0.76) | 0.40 (0.20–0.57) |
| mini-MDS | 0.66 (0.52–0.78) | 0.24 (0.14–0.40) | 0.00 (0.00–0.18) | 0.97 (0.91–0.99) | 0.00 (0.00–0.71) | 0.48 (0.46–0.50) | 0.00 (0.00–0.00) |
| Ensemble PD | 0.77 (0.64–0.91) | 0.56 (0.36–0.79) | 0.58 (0.33–0.80) | 0.89 (0.81–0.95) | 0.52 (0.30–0.74) | 0.74 (0.63–0.86) | 0.55 (0.34–0.72) |
| mini-Ensemble PD | 0.77 (0.65–0.89) | 0.45 (0.28–0.73) | 0.63 (0.38–0.84) | 0.80 (0.71–0.88) | 0.39 (0.22–0.58) | 0.72 (0.60–0.84) | 0.48 (0.29–0.64) |
| Maximum-score screening | — | — | 0.68 (0.43–0.87) | 0.77 (0.67–0.85) | 0.37 (0.21–0.55) | 0.73 (0.61–0.83) | 0.48 (0.30–0.63) |
| Sequential screening | — | — | 0.37 (0.16–0.62) | 0.97 (0.91–0.99) | 0.70 (0.35–0.93) | 0.67 (0.56–0.78) | 0.48 (0.24–0.70) |
| **Either pathological hallmark** | | | | | | | |
| SAM-PD | 0.79 (0.67–0.90) | 0.59 (0.40–0.79) | 0.59 (0.39–0.78) | 0.90 (0.82–0.95) | 0.64 (0.43–0.82) | 0.75 (0.64–0.85) | 0.62 (0.43–0.76) |
| MDS | 0.76 (0.65–0.85) | 0.50 (0.35–0.68) | 0.37 (0.19–0.58) | 0.88 (0.79–0.94) | 0.48 (0.26–0.70) | 0.62 (0.53–0.72) | 0.42 (0.23–0.57) |
| mini-MDS | 0.70 (0.59–0.80) | 0.37 (0.24–0.55) | 0.04 (0.00–0.19) | 0.98 (0.92–1.00) | 0.33 (0.01–0.91) | 0.51 (0.48–0.55) | 0.07 (0.00–0.20) |
| Ensemble PD | 0.81 (0.70–0.91) | 0.67 (0.50–0.83) | 0.52 (0.32–0.71) | 0.92 (0.84–0.97) | 0.67 (0.43–0.85) | 0.72 (0.62–0.82) | 0.58 (0.40–0.74) |
| mini-Ensemble PD | 0.80 (0.69–0.90) | 0.60 (0.41–0.79) | 0.67 (0.46–0.83) | 0.84 (0.75–0.91) | 0.56 (0.38–0.74) | 0.75 (0.65–0.85) | 0.61 (0.44–0.75) |
| Maximum-score screening | — | — | 0.67 (0.46–0.83) | 0.80 (0.70–0.88) | 0.50 (0.33–0.67) | 0.73 (0.62–0.83) | 0.57 (0.40–0.71) |
| Sequential screening | — | — | 0.30 (0.14–0.50) | 0.98 (0.92–1.00) | 0.80 (0.44–0.97) | 0.64 (0.55–0.73) | 0.43 (0.22–0.61) |
| **Both pathological hallmarks** | | | | | | | |
| SAM-PD | 0.94 (0.86–1.00) | 0.57 (0.30–0.96) | 0.86 (0.42–1.00) | 0.90 (0.82–0.95) | 0.40 (0.16–0.68) | 0.88 (0.69–0.97) | 0.55 (0.26–0.78) |
| MDS | 0.93 (0.86–0.98) | 0.56 (0.23–0.85) | 0.71 (0.29–0.96) | 0.88 (0.79–0.94) | 0.31 (0.11–0.59) | 0.80 (0.58–0.95) | 0.43 (0.16–0.67) |
| mini-MDS | 0.74 (0.58–0.87) | 0.15 (0.07–0.35) | 0.00 (0.00–0.41) | 0.98 (0.92–1.00) | 0.00 (0.00–0.84) | 0.49 (0.47–0.50) | 0.00 (0.00–0.00) |
| Ensemble PD | 0.97 (0.90–1.00) | 0.81 (0.50–1.00) | 0.86 (0.42–1.00) | 0.92 (0.84–0.97) | 0.46 (0.19–0.75) | 0.89 (0.71–0.98) | 0.60 (0.31–0.83) |
| mini-Ensemble PD | 0.94 (0.88–0.99) | 0.53 (0.28–0.92) | 0.86 (0.42–1.00) | 0.84 (0.75–0.91) | 0.30 (0.12–0.54) | 0.85 (0.66–0.95) | 0.44 (0.20–0.67) |
| Maximum-score screening | — | — | 0.86 (0.42–1.00) | 0.80 (0.70–0.88) | 0.25 (0.10–0.47) | 0.83 (0.64–0.93) | 0.39 (0.17–0.60) |
| Sequential screening | — | — | 0.71 (0.29–0.96) | 0.98 (0.92–1.00) | 0.71 (0.29–0.96) | 0.85 (0.64–0.99) | 0.71 (0.36–0.94) |

**Supplementary Table 7 |** Detailed performance metrics for all screening scores and configurations for detecting pathological hallmarks. All metrics are reported as point estimates with 95% confidence intervals (CIs); CIs for sensitivity, specificity, and precision are exact Clopper-Pearson binomial intervals and those for all other metrics were obtained from 1,000 bootstrap resamples with replacement. In the case of SAM-PD, Ensemble PD and mini-Ensemble PD scores, sensitivity, specificity, precision, balanced accuracy and F1 are reported at the Youden-optimal operating point. For the MDS and mini-MDS scores, the recommended threshold of 0.80 was used. Max-score screening and sequential screening are rule-based binary integration methods; therefore, AUROC and AUPRC are not applicable.

| **Operating point** | **AUROC** | **AUPRC** | **Sensitivity** | **Specificity** | **Precision** | **Balanced accuracy** | **F1 score** |
| --- | --- | --- | --- | --- | --- | --- | --- |
| Youden-optimal (0.78) | 0.95 (0.87–0.99) | 0.95 (0.89–0.99) | 0.85 (0.62–0.97) | 0.90 (0.68–0.99) | 0.89 (0.67–0.99) | 0.88 (0.77–0.97) | 0.87 (0.74–0.97) |
| Max F1 (0.60) | 0.95 (0.87–0.99) | 0.95 (0.89–0.99) | 0.85 (0.62–0.97) | 0.75 (0.51–0.91) | 0.77 (0.55–0.92) | 0.80 (0.67–0.92) | 0.81 (0.65–0.92) |
| Max sensitivity at specificity ≥ 0.50 (0.47) | 0.95 (0.87–0.99) | 0.95 (0.89–0.99) | 0.95 (0.75–1.00) | 0.65 (0.41–0.85) | 0.73 (0.52–0.88) | 0.80 (0.69–0.91) | 0.83 (0.70–0.93) |

**Supplementary Table 8 |** Detailed performance metrics of the SAM-PD model trained on Verily Study Watch data (PwPD n = 133, HCs n = 26) for classifying PwPD (n = 20) and HCs (n = 20) in the independent dBM-DEV cohort. All metrics are reported as point estimates with 95% confidence intervals (CIs); CIs for sensitivity, specificity, and precision are exact Clopper-Pearson binomial intervals, whereas those for all other metrics were obtained from 1,000 bootstrap resamples with replacement. Sensitivity, specificity, precision, balanced accuracy, and F1 score are reported at three operating points derived in the Verily Study Watch training dataset: the Youden-optimal point, the maximum-F1 score point, and the point that maximizes sensitivity while maintaining specificity ≥ 0.50.

| **Metric category / Feature** | **Median (IQR)** | | | **Unadjusted for age and sex (Two-sided Mann-Whitney U)** | | | **Adjusted for age and sex (Rank ANCOVA)** | | |
| --- | --- | --- | --- | --- | --- | --- | --- | --- | --- |
|  | **HC** | **At-risk** | **PwPD** | **HC**  **vs  At-risk** | **At-risk vs PwPD** | **HC  vs PwPD** | **HC  vs  At-risk** | **At-risk vs PwPD** | **HC  vs PwPD** |
| **Verily Study Watch cohort** | | | | | | | | | |
| **Sleep** | | | | | | | | | |
| Sleep efficiency (mean) | 80.1 (73.9–84.2) | 78.1 (73.4–83.8) | 74.6 (70.2–79.0) | 1.000 | < 0.001 | 0.017 | 1.000 | < 0.001 | 0.037 |
| REM sleep latency (mean) | 85.7 (72.9–117) | 102 (74.3–126) | 182 (118–292) | 1.000 | < 0.001 | < 0.001 | 0.361 | < 0.001 | < 0.001 |
| REM sleep duration (median) | 118 (70.2–149) | 110 (68.4–150) | 14.5 (0.50–53.0) | 1.000 | < 0.001 | < 0.001 | 0.172 | < 0.001 | < 0.001 |
| NREM sleep duration (median) | 261 (250–302) | 275 (234–318) | 316 (257–364) | 1.000 | 0.001 | 0.127 | 1.000 | 0.036 | 0.064 |
| **Upper-limb movement** | | | | | | | | | |
| Peak acceleration Q75 (Q75) | 0.73 (0.69–0.77) | 0.76 (0.68–0.80) | 0.60 (0.52–0.67) | 1.000 | < 0.001 | < 0.001 | 1.000 | < 0.001 | < 0.001 |
| Short-term acceleration entropy median (mean) | 2.86 (2.84–2.88) | 2.86 (2.84–2.88) | 2.91 (2.89–2.93) | 1.000 | < 0.001 | < 0.001 | 0.691 | < 0.001 | < 0.001 |
| Short-term acceleration entropy median (SD) | 0.097 (0.089–0.10) | 0.093 (0.087–0.100) | 0.083 (0.078–0.090) | 1.000 | < 0.001 | < 0.001 | 0.604 | < 0.001 | < 0.001 |
| Hourly acceleration (Q75) | 42.3 (35.8–47.6) | 38.8 (31.1–45.3) | 29.7 (24.0–37.1) | 0.496 | < 0.001 | < 0.001 | 0.046 | < 0.001 | < 0.001 |
| **Ambulatory activity** | | | | | | | | | |
| Hourly steps (mean) | 565 (421–627) | 491 (382–637) | 374 (294–474) | 0.745 | < 0.001 | < 0.001 | 0.372 | < 0.001 | < 0.001 |
| Hourly steps (SD) | 529 (391–648) | 498 (380–611) | 381 (265–520) | 1.000 | < 0.001 | 0.004 | 0.956 | 0.002 | 0.002 |
| Cadence median (Q25) | 58.5 (54.0–62.4) | 58.5 (51.0–63.0) | 45.0 (36.0–51.0) | 1.000 | < 0.001 | < 0.001 | 0.371 | < 0.001 | < 0.001 |
| Cadence SD (Q75) | 25.1 (24.0–26.1) | 25.5 (24.1–27.1) | 27.5 (25.5–29.4) | 1.000 | < 0.001 | < 0.001 | 1.000 | < 0.001 | < 0.001 |
| Cadence SD (SD) | 10.1 (9.66–10.3) | 10.2 (9.77–10.5) | 10.5 (10.1–11.0) | 1.000 | < 0.001 | 0.004 | 1.000 | < 0.001 | 0.001 |
| Cadence Q75 (SD) | 16.1 (15.1–18.4) | 16.5 (14.9–18.2) | 18.7 (17.4–20.7) | 1.000 | < 0.001 | < 0.001 | 1.000 | < 0.001 | < 0.001 |
| Walking-bout acceleration (SD) | 37.7 (31.0–46.5) | 30.0 (25.1–39.1) | 27.0 (20.6–39.3) | 0.049 | 0.316 | 0.019 | < 0.001 | 1.000 | < 0.001 |
| **dBM-DEV cohort** | | | | | | | | | |
| **Sleep** | | | | | | | | | |
| Sleep efficiency (mean) | 75.7 (69.5–79.9) | 76.7 (73.3–80.1) | 72.4 (67.7–77.6) | 1.000 | 0.187 | 0.542 | 1.000 | 1.000 | 0.630 |
| REM sleep latency (mean) | 98.7 (86.1–127) | 164 (129–236) | 141 (122–231) | 0.005 | 1.000 | 0.014 | 0.014 | 1.000 | 0.030 |
| REM sleep duration (median) | 130 (111–166) | 64.5 (10.2–80.5) | 29.5 (9.50–78.6) | < 0.001 | 1.000 | < 0.001 | 0.005 | 1.000 | < 0.001 |
| NREM sleep duration (median) | 245 (232–316) | 374 (332–424) | 327 (271–363) | < 0.001 | 0.067 | 0.098 | 0.005 | 0.575 | 0.072 |
| **Upper-limb movement** | | | | | | | | | |
| Peak acceleration Q75 (Q75) | 0.78 (0.71–0.82) | 0.73 (0.67–0.77) | 0.57 (0.55–0.65) | 0.249 | 0.002 | < 0.001 | 1.000 | < 0.001 | < 0.001 |
| Short-term acceleration entropy median (mean) | 2.86 (2.84–2.88) | 2.88 (2.86–2.89) | 2.92 (2.90–2.93) | 0.176 | < 0.001 | < 0.001 | 1.000 | < 0.001 | < 0.001 |
| Short-term acceleration entropy median (SD) | 0.093 (0.087–0.10) | 0.089 (0.086–0.095) | 0.079 (0.073–0.094) | 0.984 | 0.114 | 0.029 | 1.000 | 0.076 | 0.048 |
| Hourly acceleration (Q75) | 41.3 (35.7–51.0) | 33.8 (28.5–38.2) | 31.7 (23.7–38.3) | 0.021 | 1.000 | 0.025 | 0.384 | 0.069 | 0.005 |
| **Ambulatory activity** | | | | | | | | | |
| Hourly steps (mean) | 611 (566–771) | 580 (499–625) | 571 (451–673) | 0.362 | 1.000 | 0.542 | 1.000 | 1.000 | 0.390 |
| Hourly steps (SD) | 641 (547–746) | 626 (509–794) | 602 (504–820) | 1.000 | 1.000 | 1.000 | 1.000 | 1.000 | 1.000 |
| Cadence median (Q25) | 58.5 (54.0–66.0) | 54.0 (48.0–60.0) | 43.5 (40.7–54.0) | 0.405 | 0.009 | < 0.001 | 1.000 | 0.001 | < 0.001 |
| Cadence SD (Q75) | 23.1 (22.3–24.1) | 24.2 (21.8–25.2) | 27.5 (26.4–29.6) | 1.000 | < 0.001 | < 0.001 | 1.000 | 0.004 | < 0.001 |
| Cadence SD (SD) | 9.52 (9.38–9.95) | 9.60 (9.19–9.98) | 10.5 (10.0–11.1) | 1.000 | 0.001 | < 0.001 | 1.000 | 0.001 | < 0.001 |
| Cadence Q75 (SD) | 14.8 (13.9–15.4) | 15.7 (14.1–17.5) | 19.1 (17.1–20.3) | 0.590 | 0.001 | < 0.001 | 1.000 | < 0.001 | < 0.001 |
| Walking-bout acceleration (SD) | 32.4 (25.4–41.9) | 25.0 (20.9–31.9) | 21.3 (18.8–31.7) | 0.122 | 1.000 | 0.116 | 1.000 | 0.838 | 0.038 |

**Supplementary Table 9 |** Age- and sex-adjusted comparisons of the SAM-PD input features between groups in the Verily Study Watch and dBM-DEV cohorts. Values are median (interquartile range). Unadjusted comparisons were performed using the two-sided Mann-Whitney U test, whereas comparisons adjusted for age and sex were performed using ANCOVA on rank-transformed feature values with age and sex as covariates and heteroscedasticity-consistent (HC3) standard errors. All p values are Bonferroni-corrected for the three pairwise comparisons per feature within each cohort.

| **Metric category** | **Feature** | **Importance, mean \|SHAP\| (%)** | **ICC(2,1) Verily Study Watch, PwPD and HCs** | **ICC(2,1) dBM-DEV cohort** | **ICC(2,1) Verily Study Watch, all groups** |
| --- | --- | --- | --- | --- | --- |
| Upper-limb movement | Short-term acceleration entropy median (mean) | 15.5 | 0.95 (0.94–0.97) | 0.95 (0.91–0.97) | 0.95 (0.94–0.96) |
| Upper-limb movement | Peak acceleration Q75 (Q75) | 15.2 | 0.93 (0.90–0.95) | 0.97 (0.94–0.98) | 0.95 (0.94–0.96) |
| Upper-limb movement | Short-term acceleration entropy median (SD) | 5.3 | 0.82 (0.76–0.87) | 0.83 (0.72–0.90) | 0.81 (0.76–0.85) |
| Upper-limb movement | Hourly acceleration (Q75) | 3.9 | 0.91 (0.87–0.94) | 0.92 (0.87–0.96) | 0.91 (0.88–0.93) |
| Sleep | REM sleep duration (median) | 20.8 | 0.96 (0.94–0.97) | 0.95 (0.92–0.97) | 0.94 (0.93–0.96) |
| Sleep | REM sleep latency (mean) | 5.4 | 0.86 (0.81–0.90) | 0.91 (0.84–0.95) | 0.86 (0.82–0.89) |
| Sleep | Sleep efficiency (mean) | 4.0 | 0.83 (0.77–0.88) | 0.86 (0.76–0.92) | 0.85 (0.81–0.88) |
| Sleep | NREM sleep duration (median) | 1.2 | 0.87 (0.81–0.91) | 0.93 (0.88–0.96) | 0.86 (0.82–0.89) |
| Ambulatory activity | Cadence median (Q25) | 8.6 | 0.98 (0.97–0.99) | 0.97 (0.94–0.98) | 0.98 (0.97–0.98) |
| Ambulatory activity | Cadence SD (Q75) | 3.9 | 0.94 (0.92–0.96) | 0.93 (0.88–0.96) | 0.94 (0.92–0.96) |
| Ambulatory activity | Cadence Q75 (SD) | 3.8 | 0.82 (0.76–0.87) | 0.92 (0.86–0.95) | 0.86 (0.82–0.89) |
| Ambulatory activity | Hourly steps (mean) | 3.0 | 0.91 (0.87–0.94) | 0.83 (0.72–0.90) | 0.89 (0.86–0.92) |
| Ambulatory activity | Cadence SD (SD) | 2.6 | 0.88 (0.84–0.92) | 0.86 (0.76–0.92) | 0.85 (0.81–0.89) |
| Ambulatory activity | Hourly steps (SD) | 2.0 | 0.88 (0.83–0.91) | 0.87 (0.78–0.92) | 0.86 (0.82–0.89) |
| Ambulatory activity | Walking-bout acceleration (SD) | 1.2 | 0.88 (0.84–0.92) | 0.86 (0.76–0.92) | 0.88 (0.85–0.91) |
| Covariates | Age | 2.3 | N.A. | N.A. | N.A. |
| Covariates | Weight | 0.7 | N.A. | N.A. | N.A. |
| Covariates | Height | 0.5 | N.A. | N.A. | N.A. |
| Covariates | Sex | 0.1 | N.A. | N.A. | N.A. |
| Model output | SAM-PD score | N.A. | 0.96 (0.94–0.97) | 0.96 (0.93–0.98) | 0.96 (0.94–0.97) |

**Supplementary Table 10 |** SAM-PD model input features, feature importance, and test-retest reliability metrics. For each feature, importance is reported as its share (%) of the total mean absolute out-of-fold SHAP value (Fig. 2). Test-retest reliability is presented as the ICC(2,1) with 95% CI for sleep and activity metrics aggregated over 14 days, as well as for the resulting SAM-PD score, in the Verily Study Watch PwPD and HCs (n = 119), the independent dBM-DEV cohort (n = 50), and all Verily Study Watch groups (n = 215). N.A.: not applicable.

| **Meta-learner** | **SAM-PD score coefficient** | **MDS / mini-MDS risk score coefficient** | **Intercept** |
| --- | --- | --- | --- |
| Ensemble PD score logistic regression | 7.33 | 1.70 | -6.32 |
| mini-Ensemble PD score logistic regression | 6.12 | 2.69 | -4.98 |

**Supplementary Table 11 |** Coefficients of the logistic regression meta-learners fusing the SAM-PD and MDS scores. Both models are L2-regularized logistic regressions utilizing probabilities (0.00–1.00) as inputs without feature scaling, applying balanced class weights. The Ensemble PD score incorporates the MDS risk score and the mini-Ensemble PD score the mini-MDS risk score.

| **Operating point** | **Pathological hallmark** | **Score** | **Threshold** | **Sensitivity** | **Specificity** | **Precision** | **Balanced accuracy** | **F1 score** |
| --- | --- | --- | --- | --- | --- | --- | --- | --- |
| Max F1 | Dopaminergic deficit | SAM-PD | 0.60 | 0.80 (0.52–0.96) | 0.75 (0.65–0.82) | 0.31 (0.17–0.48) | 0.77 (0.64–0.88) | 0.44 (0.25–0.60) |
|  |  | Ensemble PD | 0.35 | 0.80 (0.52–0.96) | 0.78 (0.69–0.86) | 0.34 (0.19–0.52) | 0.79 (0.66–0.89) | 0.48 (0.28–0.63) |
|  |  | mini-Ensemble PD | 0.35 | 0.87 (0.60–0.98) | 0.75 (0.66–0.83) | 0.33 (0.19–0.50) | 0.81 (0.70–0.90) | 0.48 (0.29–0.63) |
| Max sensitivity, ≥ 0.50 specificity | Dopaminergic deficit | SAM-PD | 0.47 | 0.93 (0.68–1.00) | 0.56 (0.46–0.65) | 0.23 (0.13–0.35) | 0.74 (0.65–0.82) | 0.37 (0.21–0.50) |
|  |  | Ensemble PD | 0.12 | 0.93 (0.68–1.00) | 0.65 (0.55–0.74) | 0.27 (0.16–0.42) | 0.79 (0.70–0.86) | 0.42 (0.26–0.56) |
|  |  | mini-Ensemble PD | 0.23 | 0.93 (0.68–1.00) | 0.70 (0.60–0.78) | 0.30 (0.18–0.46) | 0.82 (0.72–0.88) | 0.46 (0.29–0.60) |
| Max F1 | α-Synuclein pathology | SAM-PD | 0.60 | 0.63 (0.38–0.84) | 0.73 (0.63–0.81) | 0.32 (0.18–0.49) | 0.68 (0.56–0.80) | 0.42 (0.25–0.58) |
|  |  | Ensemble PD | 0.35 | 0.63 (0.38–0.84) | 0.77 (0.67–0.85) | 0.35 (0.20–0.54) | 0.70 (0.58–0.82) | 0.45 (0.27–0.61) |
|  |  | mini-Ensemble PD | 0.35 | 0.68 (0.43–0.87) | 0.74 (0.64–0.82) | 0.34 (0.20–0.51) | 0.71 (0.60–0.83) | 0.46 (0.28–0.61) |
| Max sensitivity, ≥ 0.50 specificity | α-Synuclein pathology | SAM-PD | 0.47 | 0.79 (0.54–0.94) | 0.54 (0.43–0.64) | 0.25 (0.15–0.38) | 0.66 (0.56–0.76) | 0.38 (0.24–0.51) |
|  |  | Ensemble PD | 0.12 | 0.79 (0.54–0.94) | 0.65 (0.55–0.75) | 0.31 (0.19–0.46) | 0.72 (0.62–0.82) | 0.45 (0.29–0.59) |
|  |  | mini-Ensemble PD | 0.23 | 0.68 (0.43–0.87) | 0.67 (0.57–0.77) | 0.30 (0.17–0.45) | 0.68 (0.57–0.80) | 0.41 (0.25–0.56) |
| Max F1 | Either pathological hallmark | SAM-PD | 0.60 | 0.67 (0.46–0.83) | 0.76 (0.66–0.85) | 0.46 (0.30–0.63) | 0.72 (0.61–0.82) | 0.55 (0.38–0.69) |
|  |  | Ensemble PD | 0.35 | 0.67 (0.46–0.83) | 0.81 (0.71–0.88) | 0.51 (0.34–0.69) | 0.74 (0.63–0.84) | 0.58 (0.41–0.71) |
|  |  | mini-Ensemble PD | 0.35 | 0.70 (0.50–0.86) | 0.78 (0.67–0.86) | 0.49 (0.32–0.65) | 0.74 (0.64–0.84) | 0.58 (0.41–0.72) |
| Max sensitivity, ≥ 0.50 specificity | Either pathological hallmark | SAM-PD | 0.47 | 0.81 (0.62–0.94) | 0.56 (0.45–0.67) | 0.36 (0.24–0.49) | 0.69 (0.59–0.77) | 0.50 (0.36–0.62) |
|  |  | Ensemble PD | 0.12 | 0.81 (0.62–0.94) | 0.69 (0.58–0.78) | 0.44 (0.30–0.59) | 0.75 (0.65–0.83) | 0.57 (0.42–0.70) |
|  |  | mini-Ensemble PD | 0.23 | 0.74 (0.54–0.89) | 0.71 (0.60–0.80) | 0.43 (0.29–0.59) | 0.72 (0.62–0.82) | 0.55 (0.38–0.68) |
| Max F1 | Both pathological hallmarks | SAM-PD | 0.60 | 0.86 (0.42–1.00) | 0.76 (0.66–0.85) | 0.22 (0.09–0.42) | 0.81 (0.62–0.92) | 0.35 (0.15–0.55) |
|  |  | Ensemble PD | 0.35 | 0.86 (0.42–1.00) | 0.81 (0.71–0.88) | 0.26 (0.10–0.48) | 0.83 (0.65–0.94) | 0.40 (0.17–0.62) |
|  |  | mini-Ensemble PD | 0.35 | 1.00 (0.59–1.00) | 0.78 (0.67–0.86) | 0.26 (0.11–0.46) | 0.89 (0.84–0.93) | 0.41 (0.20–0.62) |
| Max sensitivity, ≥ 0.50 specificity | Both pathological hallmarks | SAM-PD | 0.47 | 1.00 (0.59–1.00) | 0.56 (0.45–0.67) | 0.15 (0.06–0.29) | 0.78 (0.73–0.83) | 0.26 (0.12–0.42) |
|  |  | Ensemble PD | 0.12 | 1.00 (0.59–1.00) | 0.69 (0.58–0.78) | 0.20 (0.08–0.37) | 0.84 (0.79–0.89) | 0.33 (0.16–0.51) |
|  |  | mini-Ensemble PD | 0.23 | 1.00 (0.59–1.00) | 0.71 (0.60–0.80) | 0.21 (0.09–0.39) | 0.85 (0.80–0.90) | 0.35 (0.17–0.54) |

**Supplementary Table 12 |** Performance of the SAM-PD, Ensemble PD and mini-Ensemble PD scores for detecting pathological hallmarks in the Verily Study Watch at-risk group at two additional pre-specified operating points: the maximum-F1 score point (Max F1) and the point maximizing sensitivity with specificity ≥ 0.50 (Max sensitivity, ≥ 0.50 specificity), each derived from the PwPD versus HC classification task in the Verily Study Watch cohort. All metrics are reported as point estimates with 95% confidence intervals (CIs); CIs for sensitivity, specificity, and precision are exact Clopper-Pearson binomial intervals, whereas those for all other metrics were obtained from 1,000 bootstrap resamples with replacement.

| **Classifier** | **Hyperparameter** | **Values searched** | **Configurations** | **Selected** |
| --- | --- | --- | --- | --- |
| Balanced random forest | Number of trees | 200, 400 | 16 | 400 |
|  | Maximum depth | 4, 8 |  | 4 |
|  | Maximum features | 0.5, sqrt |  | sqrt |
|  | Minimum samples per leaf | 1, 5 |  | 1 |
| Ridge logistic regression | Regularization strength (C) | 0.01, 0.03, 0.1, 0.3, 1.0 | 5 | — |
|  | Penalty | L2 |  | — |
|  | Class weight | Balanced |  | — |
| Gradient-boosted trees (LightGBM) | Number of trees | 200, 400 | 8 | — |
|  | Maximum depth | 2, 4 |  | — |
|  | Learning rate | 0.05, 0.1 |  | — |
|  | Number of leaves | 31 |  | — |
|  | Class weight | Balanced |  | — |

**Supplementary Table 13** | Hyperparameter search grids for the candidate classifiers. The final column indicates the balanced random forest configuration selected for the final SAM-PD model.

| **Category** | **Variable** | **PPMI source or reason for exclusion** | **MDS risk score** | **mini-MDS risk score** |
| --- | --- | --- | --- | --- |
| Demographics | Biological sex | Sex (Male / Female) | ✓ | ✓ |
| Lifestyle Factors | Never smoked | "Have you ever smoked regularly?" = No | ✓ | ✓ |
|  | Current smoker | "Are you a current smoker?" = Yes | ✓ | ✓ |
|  | Former smoker | Ever smoked + Not current smoker | ✓ | ✓ |
|  | Caffeine consumption | Drank caffeinated coffee regularly | ✓ | ✓ |
|  | Physical inactivity | *No relevant assessment or measure in PPMI* |  |  |
|  | Occupational pesticide exposure | Job exposure to pesticides | ✓ |  |
|  | Occupational solvent exposure | *Not systematically collected in PPMI* |  |  |
| Genetic Risk Factors | Polygenic risk score | High / Low percentile of AMP-PD PRS | ✓ |  |
|  | GBA mutation present | From Participant Status | ✓ |  |
|  | LRRK2 mutation present | From Participant Status | ✓ |  |
|  | First-degree relative with PD | From Family History | ✓ | ✓ |
| Medical Conditions | Diabetes mellitus (Type II) | Medication Log records | ✓ | ✓ |
| Laboratory | Low plasma urate (males) | *Not systematically collected in PPMI* |  |  |
| Sleep Disorders | PSG-confirmed RBD | In RBD prodromal sub-cohort | ✓ |  |
|  | RBD screening questionnaire | RBDSQ ≥ 5 | ✓ | ✓ |
| Motor Features | Subthreshold Parkinsonism | MDS-UPDRS Part III > 6 (excluding postural and action tremor) | ✓ |  |
|  | Abnormal quantitative motor testing | *Alternative assessment of subthreshold parkinsonism; not used as MDS-UPDRS Part III was used instead* |  |  |
| Sensory Features | Hyposmia | From Participant Status | ✓ |  |
| Imaging | Substantia nigra hyperechogenicity | *Not systematically collected in PPMI* |  |  |
|  | Dopaminergic DAT scan | *Excluded because it was used as ground truth in this study* |  |  |
| Autonomic Features | Constipation | MDS-UPDRS Part I & Medication Log | ✓ | ✓ |
|  | Daytime somnolence | MDS-UPDRS Part I & Medication Log | ✓ | ✓ |
|  | Neurogenic orthostatic hypotension | HR / SBP ratio calculation | ✓ |  |
|  | Symptomatic orthostatic hypotension | MDS-UPDRS Part I & Medication Log | ✓ |  |
|  | Erectile dysfunction | SCOPA-AUT & Medication Log (males) | ✓ |  |
|  | Urinary dysfunction | MDS-UPDRS Part I & Medication Log | ✓ | ✓ |
| Neuropsychiatric Features | Depression | MDS-UPDRS Part I & Medication Log | ✓ | ✓ |
|  | Cognitive impairment | MDS-UPDRS Part I & Medication Log | ✓ | ✓ |

**Supplementary Table 14 |** Risk and prodromal markers according to the MDS research criteria for prodromal PD, their PPMI source or reason for exclusion, and inclusion in the calculation of the MDS and mini-MDS risk scores. Smoking status is represented by three mutually exclusive states, whereas family history, gene mutations and the polygenic risk score (PRS) are subcomponents of the genetic marker. The MDS risk score operationalized the risk and prodromal markers that were available in the PPMI dataset. The mini-MDS risk score is a version relying only on markers that do not require specialized ancillary testing and can be self-reported with relative confidence.

### **Supplementary Figures**

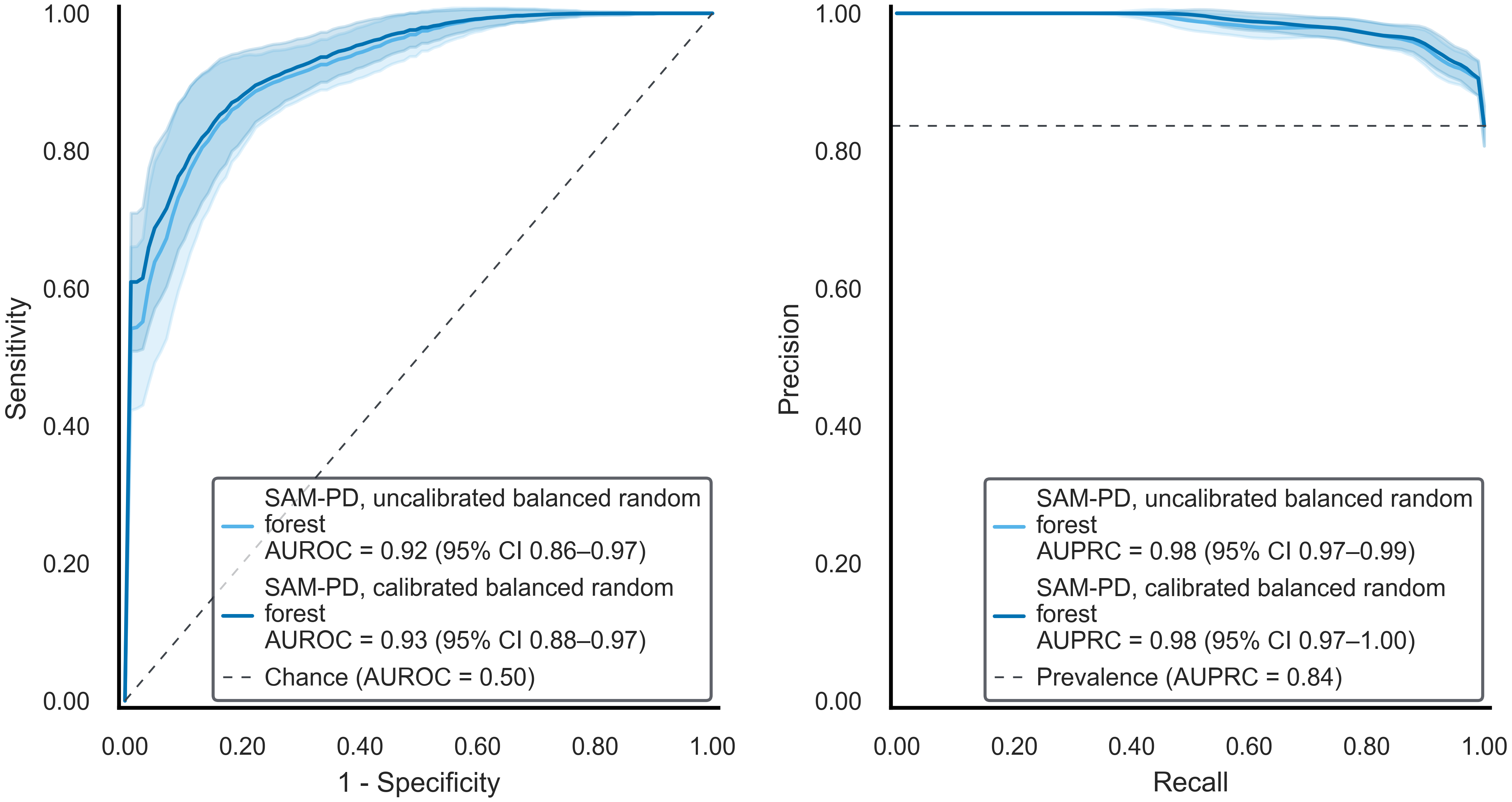

**Supplementary Fig. 1 |** SAM-PD model performance for PwPD versus HC classification in the Verily Study Watch cohort. (Left) Receiver Operating Characteristic (ROC) and (right) Precision-Recall curves compare the performance of uncalibrated and calibrated SAM-PD balanced random forest models under Leave-One-Subject-Out cross-validation. In each panel, shaded areas indicate ± 1 standard deviation of the bootstrap curves and the dashed line denotes the chance level; the inset displays the AUROC or AUPRC (95% CI) of each model, alongside the chance AUROC (0.50) or chance AUPRC (positive class prevalence).

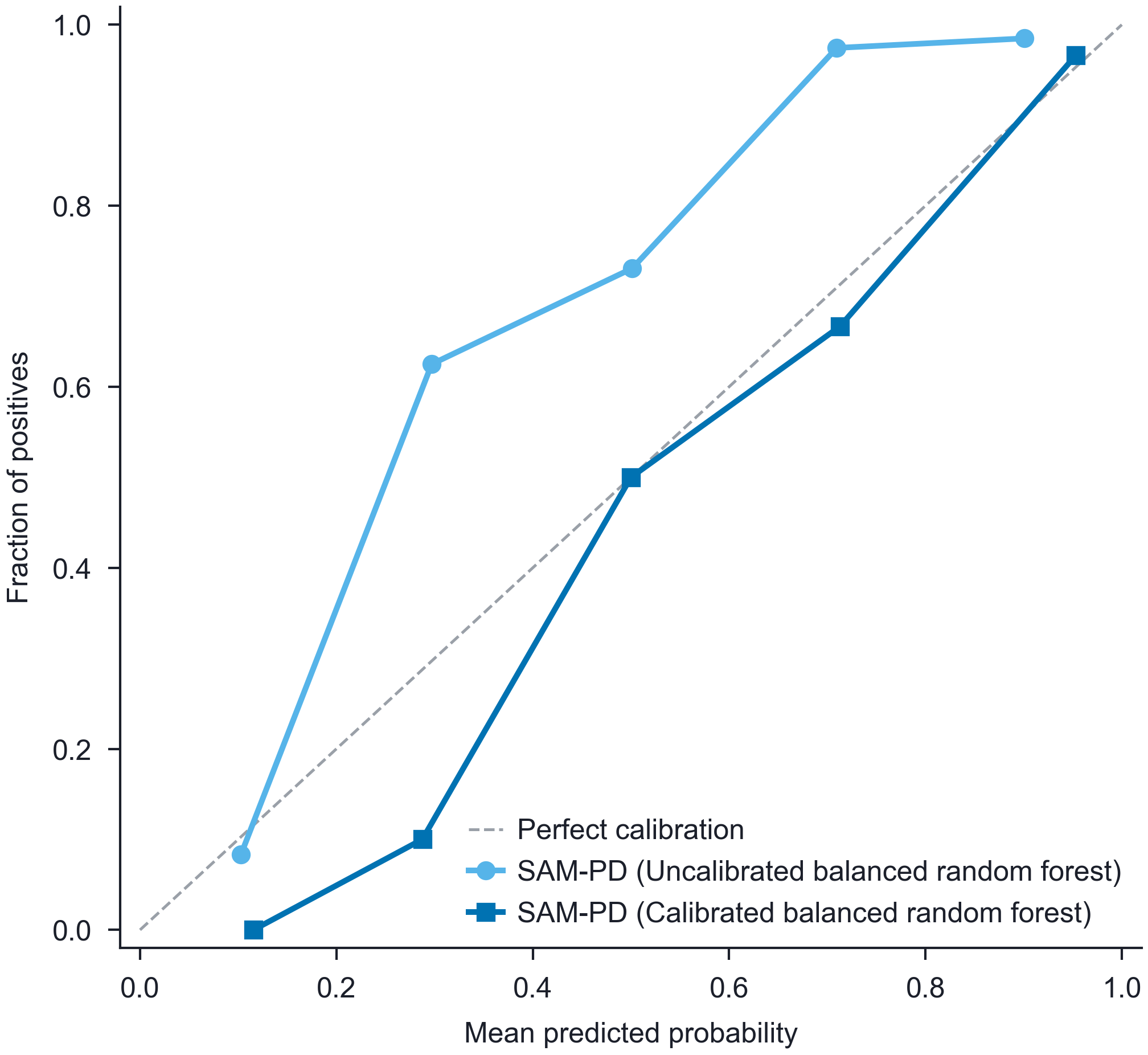

**Supplementary Fig. 2 |** Calibration curves for SAM-PD models in the Verily Study Watch cohort. Predicted probabilities are plotted against observed outcome frequencies for the uncalibrated and the calibrated balanced random forest models, evaluated under Leave-One-Subject-Out cross-validation for PwPD versus HC classification. The dashed line denotes perfect calibration.

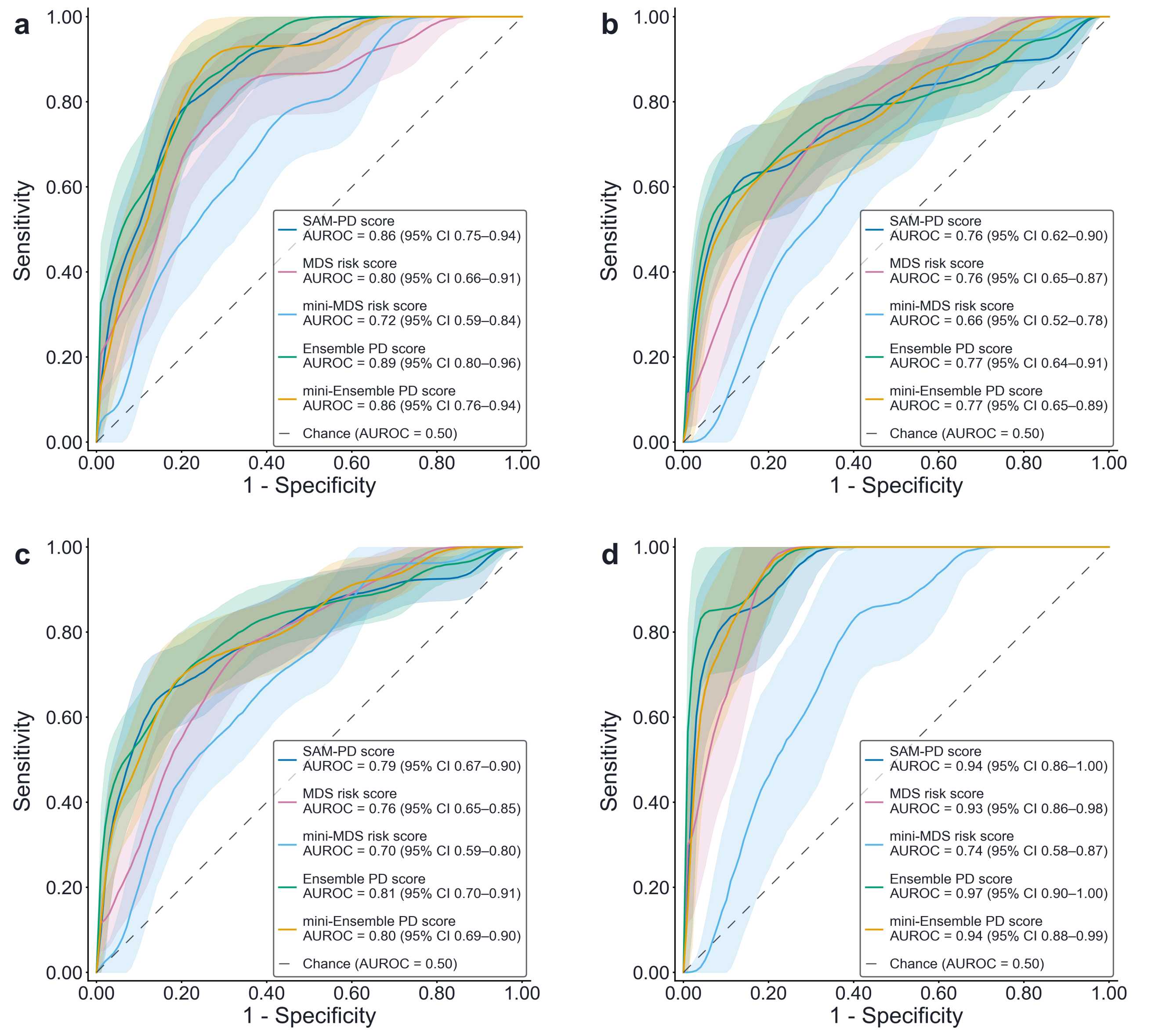

**Supplementary Fig. 3** **|** ROC-based performance for detecting PD pathological hallmarks in the Verily Study Watch at-risk group. ROC curves compare the performance of the SAM-PD, MDS risk, mini-MDS risk, Ensemble PD, and mini-Ensemble PD scores across four detection scenarios: (a) dopaminergic deficit, (b) α-synuclein pathology, (c) either pathological hallmark present, and (d) both pathological hallmarks present. In each panel, shaded areas indicate ± 1 standard deviation of the bootstrap curves and the dashed line denotes the chance level; the inset displays the AUROC (95% CI) of each model, alongside the chance AUROC (0.50).

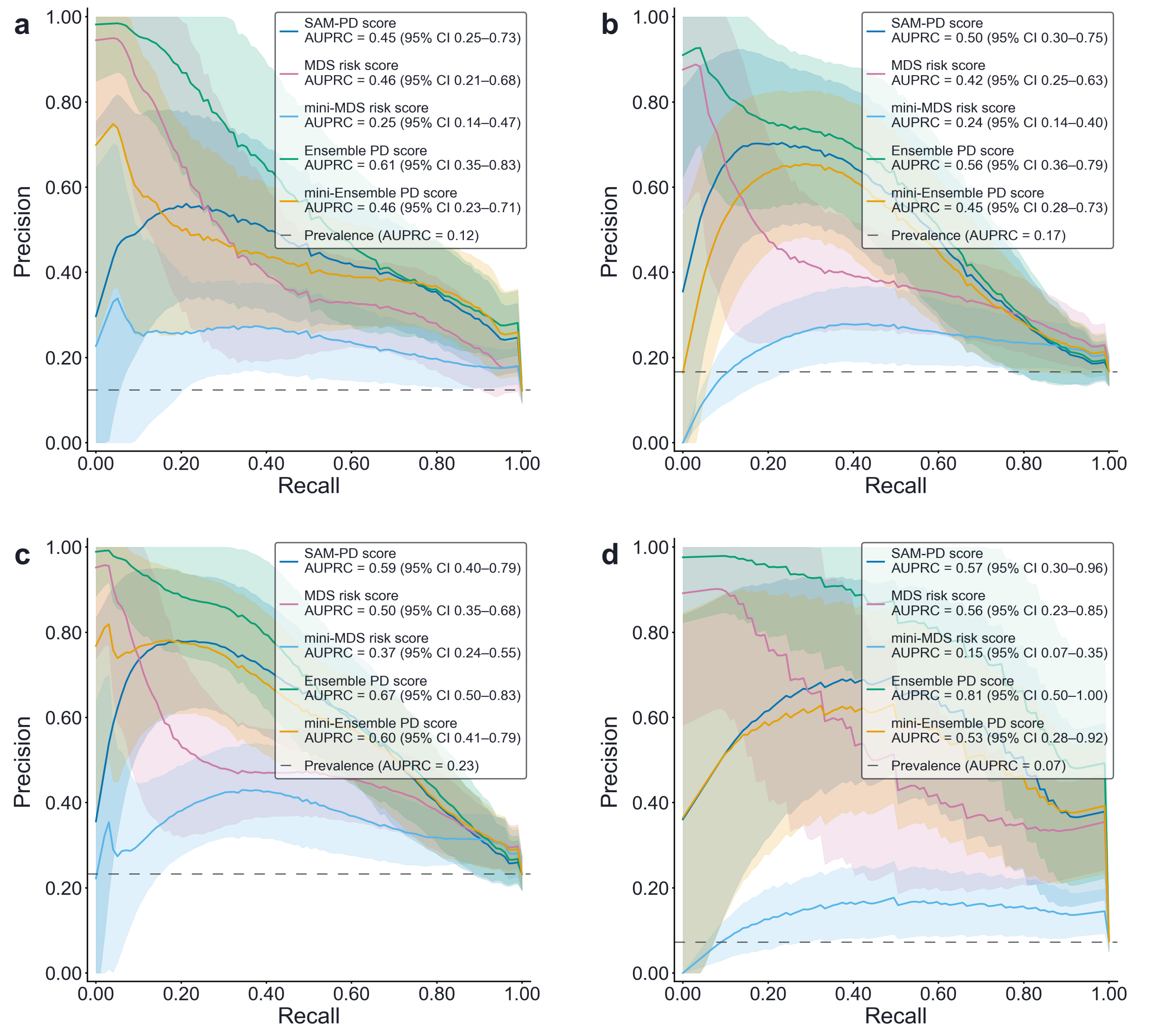

**Supplementary Fig. 4** **|** Precision-Recall-based performance for detecting PD pathological hallmarks in the Verily Study Watch at-risk group. Precision-Recall curves compare the performance of the SAM-PD, MDS risk, mini-MDS risk, Ensemble PD, and mini-Ensemble PD scores across four detection scenarios: (a) dopaminergic deficit, (b) α-synuclein pathology, (c) either pathological hallmark present, and (d) both pathological hallmarks present. In each panel, shaded areas indicate ± 1 standard deviation of the bootstrap curves and the dashed line denotes the chance level; insets report the AUPRC (95% CI) of each model, alongside the chance AUPRC (based on positive class prevalence).

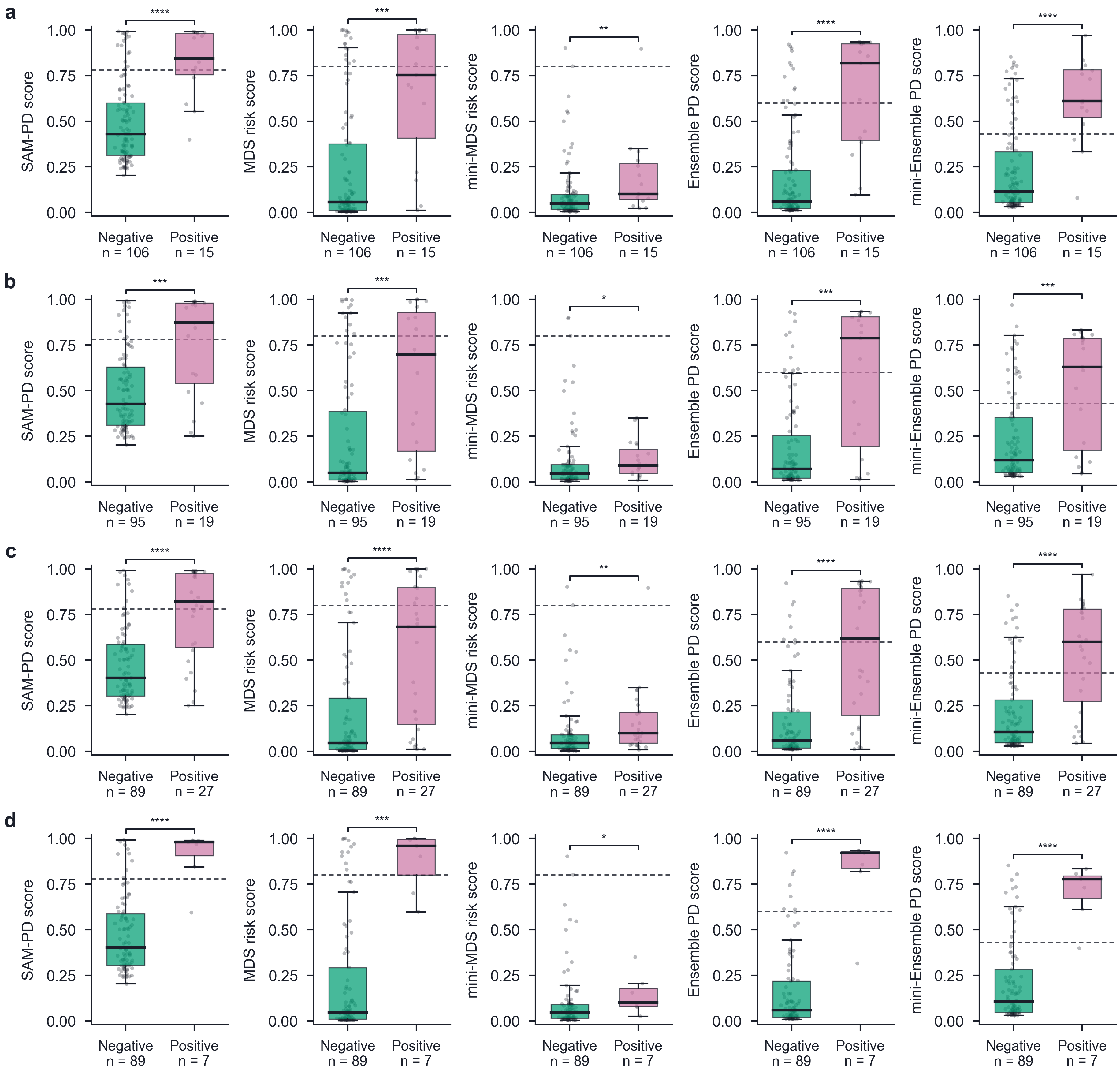

**Supplementary Fig. 5 |** Score distributions across PD pathological hallmark scenarios in the Verily Study Watch at-risk group. Box plots show the score distributions for SAM-PD, MDS risk, mini-MDS risk, Ensemble PD, and mini-Ensemble PD (left to right columns) in hallmark-negative and hallmark-positive participants across four scenarios: (a) dopaminergic deficit, (b) α-synuclein pathology, (c) either pathological hallmark present, and (d) both pathological hallmarks present. Boxes denote the interquartile range (IQR) with the median indicated by the horizontal line within each box; whiskers extend to 1.5 × IQR, and overlaid grey points represent individual participant values. Dashed lines indicate each score's operating threshold (Youden-optimal for the SAM-PD, Ensemble PD and mini-Ensemble PD scores, derived in PwPD versus HCs; 0.80 for the MDS and mini-MDS risk scores). Pairwise statistically significant differences, determined using the two-sided Mann-Whitney U test, are annotated (*, p < 0.05; **, p < 0.01; ***, p < 0.001; ****, p < 0.0001).

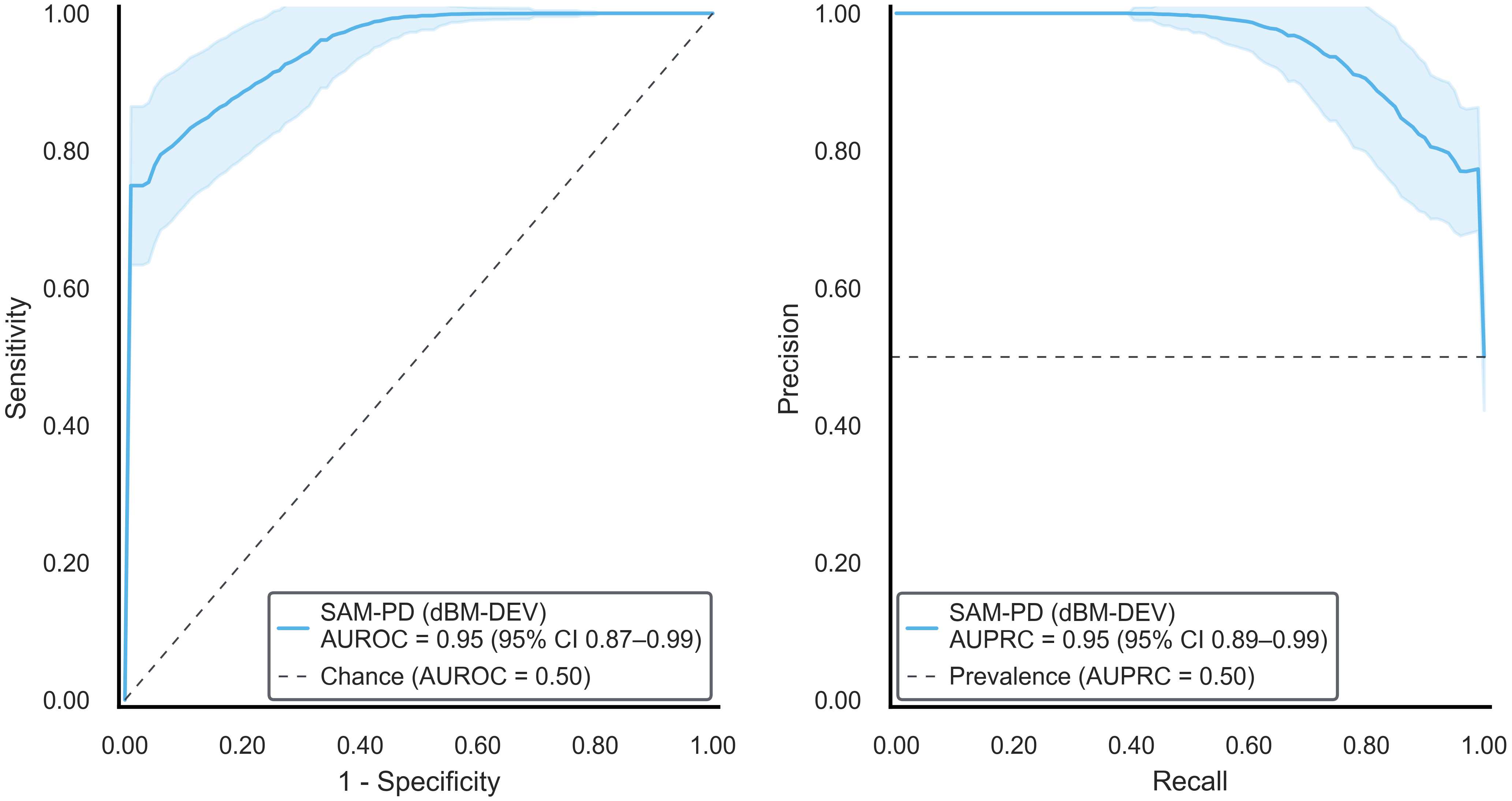

**Supplementary Fig. 6 |** SAM-PD model performance for PwPD versus HC classification in the dBM-DEV cohort. (Left) Receiver Operating Characteristic (ROC) and (right) Precision-Recall curves obtained by applying the SAM-PD model trained on Verily Study Watch data to PwPD and HCs in the independent dBM-DEV cohort. In each panel, the shaded area indicates ± 1 standard deviation of the bootstrap curves and the dashed line denotes the chance level; the inset displays the AUROC or AUPRC (95% CI) for the model, alongside the chance AUROC (0.50) or chance AUPRC (positive class prevalence).

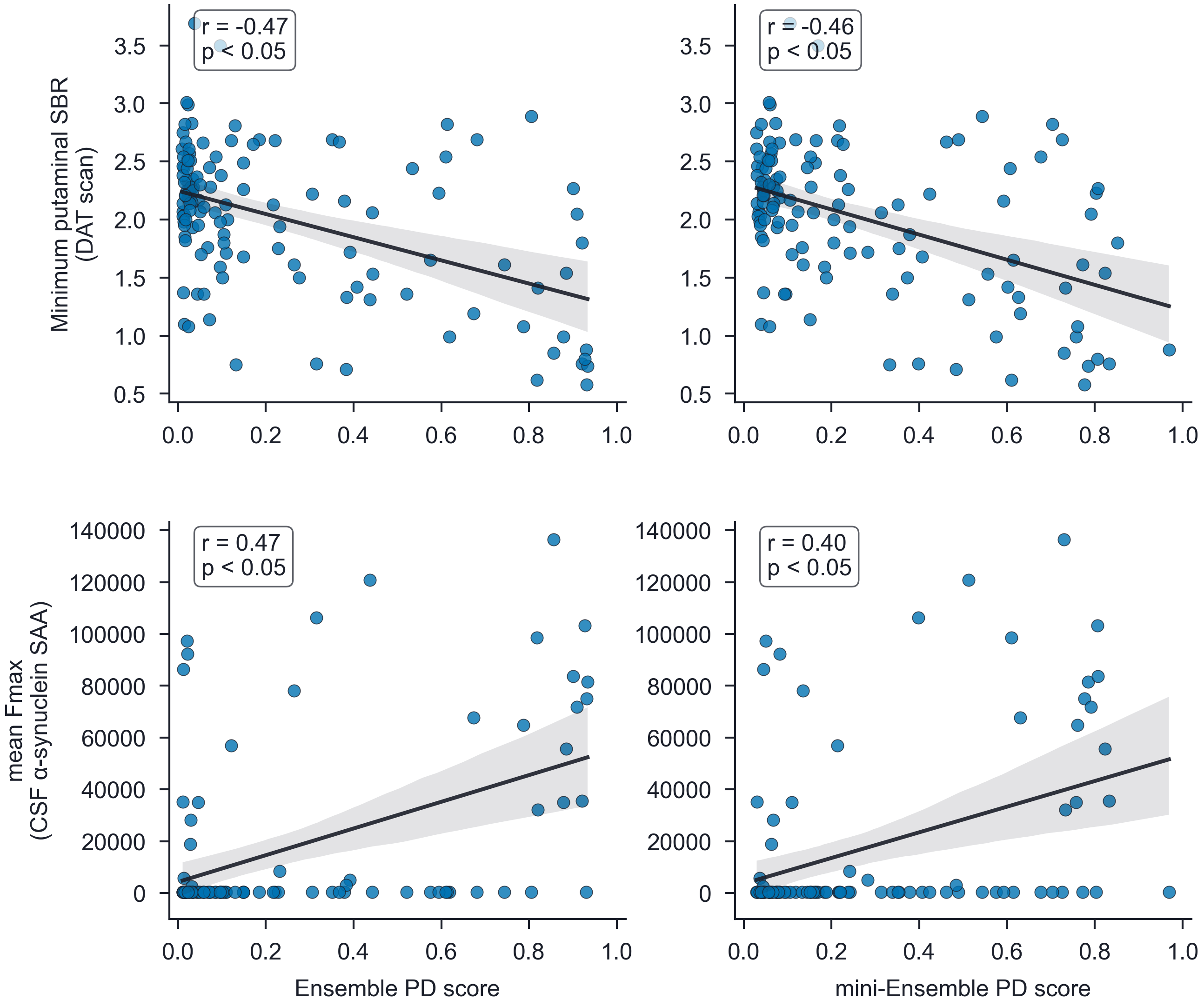

**Supplementary Fig. 7 |** Relationship of the Ensemble PD and mini-Ensemble PD scores with pathological markers of dopaminergic deficit and α-synuclein pathology in the Verily Study Watch at-risk group. Scatter plots show the Ensemble PD score (left column) and the mini-Ensemble PD score (right column) against the DAT scan-derived minimum putaminal SBR (top row) and the CSF α-synuclein SAA mean Fmax (bottom row). Mean Fmax is the mean maximum fluorescence across three SAA replicates. In each panel, the least-squares regression line and its 95% CI (shaded area) are shown; the inset displays the Pearson correlation coefficient (r) and the corresponding p-value.
